# Impact of vaccination on the speed of antigenic evolution

**DOI:** 10.64898/2026.03.10.26347605

**Authors:** Myrthe S Willemsen, Ganna Rozhnova

## Abstract

Rapidly evolving pathogens can escape antibody-mediated immunity, leading to recurrent epidemics. Vaccination is a key intervention to reduce infections and severe disease, yet concerns remain that it may accelerate antigenic evolution, potentially undermining long-term vaccine effectiveness. We developed a multi-strain mathematical model, parameterized for a rapidly evolving pathogen, to systematically explore how vaccination influences both the speed of antigenic evolution and the incidence of infection across a range of biological vaccine characteristics (efficacy, neutralization breadth, and vaccine strain) and implementation strategies (vaccination coverage and frequency). In the model, pathogen evolution is driven by cross-immunity and stochastic mutations in a one-dimensional antigenic space, and vaccination reduces an individual’s susceptibility to circulating strains according to the cross-immunity conferred by the vaccine strain. We find that vaccination generally reduces infection incidence, with higher coverage and efficacy leading to larger declines and, eventually, pathogen extinction. When transmission is substantially suppressed, antigenic evolution slows down. However, when vaccines match circulating strains but confer narrow cross-immunity, vaccination may accelerate antigenic evolution and potentially increase incidence. In a case study of seasonal influenza, vaccines with increased efficacy can speed up antigenic evolution but do not raise incidence. Overall, our results show that vaccination can effectively reduce both infection incidence and the speed of antigenic evolution in many scenarios. Nevertheless, the potential for vaccine-driven evolution warrants careful consideration, particularly when vaccine effectiveness in reducing incidence is limited.

## 1. Introduction

Many pathogens engage in a continuous evolutionary arms race with their hosts, driven by selective pressure imposed by the immune system. Through rapid evolution, they can evade immune responses, such as antibody-mediated immunity [1, 2, 3, 4]. Consequently, individuals generate new immune responses, resulting in dynamic patterns of population-level immunity. The ongoing antigenic evolution of rapidly evolving pathogens promotes re-infection and recurrent epidemics, leading to a substantial global health burden each year. At the molecular level, random mutations in key antigenic sites can reduce the effectiveness of pre-existing immune responses [5, 3]. When a pathogen is widespread, multiple advantageous strains may arise and compete for susceptible individuals, with those possessing the highest fitness advantage more likely to become established [6].

Vaccination is a critical tool for reducing the health burden caused by pathogens, including severe outcomes such as hospitalization and death. When implemented at scale, vaccination programmes can also influence pathogen evolution by reducing the pool of susceptible individuals, limiting transmission, and altering selection pressure at the population level [7]. Examples include respiratory syncytial virus [8], SARS-CoV-2 [9], and seasonal influenza [3, 10]. However, concerns have been raised that vaccination could, under certain circumstances, accelerate antigenic evolution [11, 2, 12], potentially undermining long-term vaccine effectiveness. Understanding whether vaccination might shape pathogen evolution is therefore crucial for the design of effective vaccination strategies. This question has gained particular importance following recent advances in vaccine development after the COVID-19 pandemic, which has marked a new era in vaccine design [13, 11, 12, 14].

Signs of vaccine-induced antigenic evolution have been reported for some pathogens [15, 16] (Figure 1). However, the evidence remains limited, as evolutionary trajectories are shaped by complex and overlapping processes. Ecological and epidemiological interactions, as well as non-pharmaceutical interventions, may confound efforts to link antigenic changes to vaccination, particularly for rapidly evolving pathogens [7, 17, 18]. Theoretically, two opposing hypotheses describe how vaccination may influence antigenic evolution [19, 2, 20]. On one hand, vaccination can increase the transmission advantage of immune escape strains, accelerating their establishment, similarly to the evolution of antiviral drug resistance [21]. Vaccine-induced selection has been suggested in contexts where multiple strains coexisted before vaccine introduction, such as serotype replacement after vaccination against *Streptococcus pneumoniae* [22], *Bordetella pertussis* [23], and *Haemophilus influenzae* type b [24]. For viruses, the potential for vaccine-induced evolution has been observed in animal studies, with escape mutations and increased viral diversity in vaccinated populations [25, 26]. On the other hand, by reducing the reproduction number of the pathogen and the overall number of infections, vaccination can decrease its effective population size, potentially slowing antigenic evolution [27].

**Figure 1.**
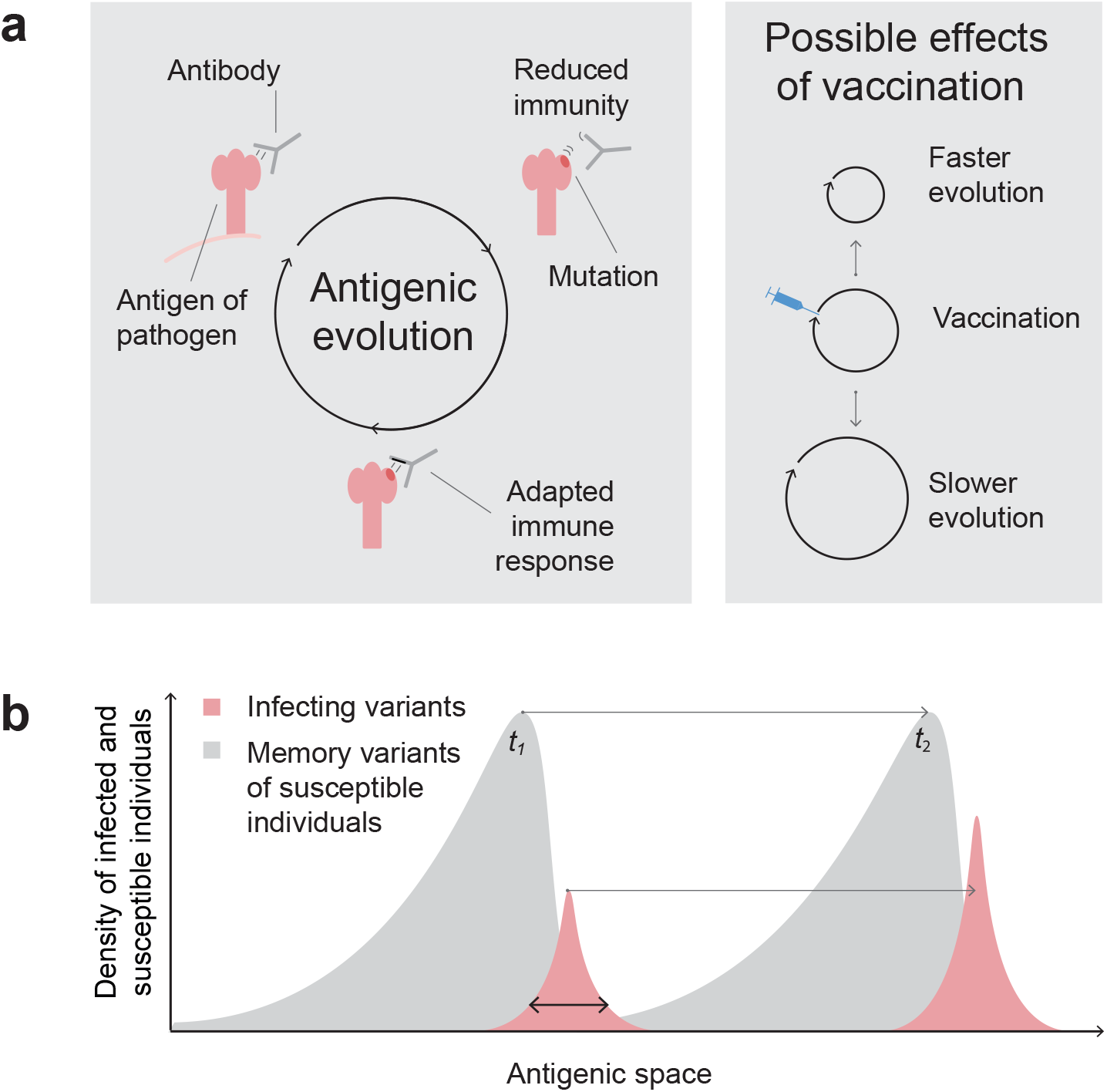
Antigenic evolution of a rapidly evolving pathogen. (**a**) Frequent mutations in regions targeted by antibodies can reduce antibody binding, increasing susceptibility to infection with mutated strains. Over time, immune responses are updated, and population-level immunity co-evolves with the pathogen. The possible effects of vaccination on the speed of antigenic evolution remain debated. (**b**) Antigenic properties of circulating strains can be represented along a one-dimensional antigenic axis. The red wave shows the density of individuals currently infected with each strain, moving through antigenic space at a characteristic speed. The horizontal arrow represents the shift in time. The red wave’s width reflects antigenic diversity. A second (grey) wave follows, representing individuals recovered from some strains and susceptible according to their immune memory. New strains arise at the leading edge of the red wave via mutation.

Previous modeling studies have reached mixed conclusions regarding vaccination’s impact on evolution. Reported conditions under which vaccination against seasonal influenza and SARS-CoV-2 may promote the emergence or establishment of vaccine escape strains vary widely [28, 19, 29, 30, 31, 32, 17]. These contrasting findings highlight the need for a generalized, pathogen-agnostic, systematic modeling exploration of the evolutionary consequences of vaccination to clarify the conditions under which vaccination accelerates or slows antigenic change.

This study aimed to understand how vaccination influences both the speed of antigenic evolution and infection incidence for rapidly evolving pathogens. Building on our previous modeling framework [33] and related approaches [34, 20], we extended the classic susceptible–infected–recovered model to incorporate vaccination and multiple antigenically distinct strains arranged in a one-dimensional antigenic space, representing the spindly, ladder-like phylogenies characteristic of seasonal influenza and coronaviruses. We systematically evaluated how vaccination strategies and vaccine characteristics shape the epidemiological and evolutionary outcomes by varying key parameters of vaccine deployment and design, including coverage, deployment frequency, efficacy, breadth, and strain composition. By exploring a broad parameter space, we identify the conditions under which vaccination may accelerate, slow, or has little effect on antigenic evolution, thereby providing a mechanistic understanding of vaccine-driven dynamics for rapidly evolving pathogens.

## 2. Methods

### 2.1. Overview

We developed a stochastic, multi-strain, population-level compartmental model to describe both transmission and antigenic evolution (hereafter, ‘evolution’) of a rapidly evolving pathogen. The model exhibits a characteristic ‘traveling wave’ dynamic in antigenic space, whereby newly emerging strains gradually replace older strains along a one-dimensional antigenic axis [35, 36]. Each strain is defined by its coordinate *x* in this antigenic space, with larger values corresponding to more recently evolved strains, *x > y*. Throughout, we use the term ‘strains’ to refer to pathogen variants distinguished by their antigenic coordinate. The model was parameterized using values representative of a generic rapidly evolving pathogen and, in a separate case study, seasonal influenza A/H3N2. The main model outcomes were the speed of antigenic evolution and infection incidence.

### 2.2. Transmission model

The transmission model was based on the susceptible-infected-recovered epidemiological dynamics, describing the evolution of the density of individuals who are currently infected with strain *x, i*(*x, t*), susceptible unvaccinated, *s*(*x, t*), and susceptible vaccinated, *v*(*x, t*). The latter two groups of individuals are both currently uninfected, who recovered from strain *x* and are currently susceptible to other strains according to their vaccine status and infection memory. The ‘infection memory’ *x* denotes an individual’s last infecting strain. Older immune memories are neglected [33]. The only group of individuals who are currently infected is *i*(*x, t*).

The dynamics of the model are governed by several processes (Figure S1). Individuals who are susceptible unvaccinated, *s*(*x, t*), and susceptible vaccinated, *v*(*x, t*), get infected with a transmission rate *β* = *R*_0_*r*, where *R*_0_ is the basic reproduction number and *r* is the recovery rate. Prior infection memory of susceptible unvaccinated, *s*(*x, t*), and susceptible vaccinated, *v*(*x, t*), individuals recovered from strain *x* reduces their susceptibility to other strains *y*, which is described by the cross-immunity functions, *K*_*s*_(*x − y*) and *K*_*v*_(*x − y, z*(*t*) *− y*), respectively. Note that while the susceptibility of unvaccinated individuals depends on cross-immunity between the infecting strain *y* and the infection memory strain *x, K*_*s*_(*x − y*), the susceptibility of vaccinated individuals also depends on cross-immunity between strain *y* and the vaccine strain *z*(*t*), *K*_*v*_(*x − y, z*(*t*) *− y*). The cross-immunity functions and the selection of the vaccine strain *z*(*t*) are described in Section 2.4. The individuals currently infected with strain *x, i*(*x, t*), recover and become susceptible, *s*(*x, t*), with a recovery rate, *r*.

Vaccination is allocated at random to a proportion *c* of the susceptible population at regular time intervals, *t*_*v*_, where *c* denotes vaccination coverage. Individuals retain immunological memory of the most recent vaccination event only. Upon receipt of a new vaccine, individuals previously vaccinated are moved back to the susceptible compartment according to their infection memories. This modeling assumption implies short-lived vaccine-induced protection. Specifically, vaccine-induced immunity wanes at a rate *γ*_*z*_, after which individuals return to the susceptible class while retaining memory of their most recent natural infection with strain *x*. Waning of natural immunity is assumed to be slower than waning of vaccine-induced immunity [37, 38], and is therefore not modeled explicitly, but replaced through new infections.

Without mutation, the dynamics of the model are described by the following set of equations

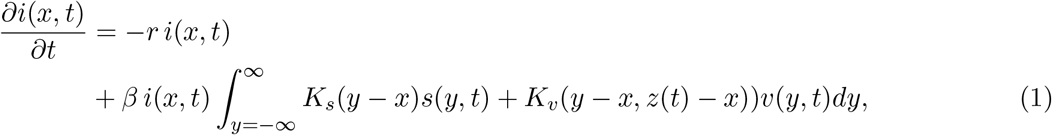

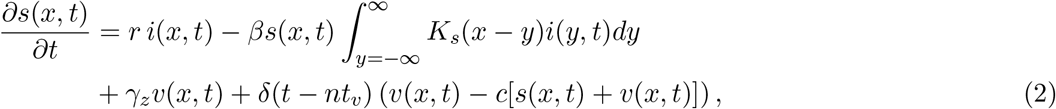

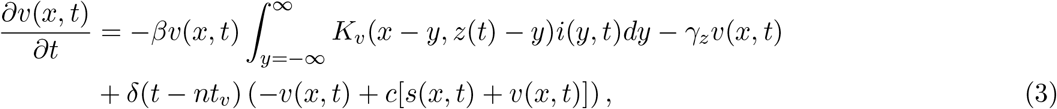

where *n* = 1, 2, 3, … indexes the vaccination events and *δ* represents the Dirac delta function. Since each individual is either currently infected, susceptible unvaccinated, or susceptible vaccinated, and the population size is conserved, we have the following normalization condition

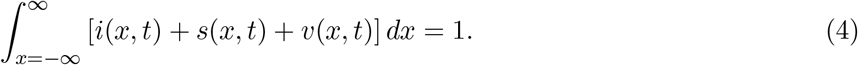

Initial and boundary conditions are discussed in the Supplement 1.1.

### 2.3. Genetic drift and mutations

Stochasticity in the model is introduced through random mutations and genetic drift. To account for genetic drift, which describes the change in the frequency of a strain due to random events [39, 40], we assumed a fixed population size *N* and stochastically simulated new infections and recoveries using a Poisson distribution.

A rapidly evolving pathogen is assumed to evolve in a one-dimensional antigenic space, representing the genealogical trunk of the phylogenetic tree. Distances between strains are calculated as the difference between their coordinates on this one-dimensional antigenic axis, expressed in ‘antigenic units’. New strains emerge via genetic mutations, particularly those that modify, to varying degrees, the pathogen’s antigenic coordinate. The mutation rate, *U*_*b*_, denotes the average number of antigenic mutations per antibody–binding region per day, under the assumption of negligible cost to the pathogen’s intrinsic fitness [41, 42]. The number of mutants generated by each strain is drawn from a Poisson distribution. For each mutant, the antigenic effect of the mutation ∆*x* is fixed at one or drawn from a probability distribution *P* (∆*x*), which we vary between gamma, half-gaussian, and exponential. Supplements 1.2 and 1.3 provide further details on genetic drift and mutations.

### 2.4. Cross-immunity

Prior infection or vaccination with one strain often confers cross-immunity, also termed cross-protection or cross-reactivity, to other antigenically similar strains. The natural cross-immunity function, *K*_*s*_(*x − y*), describes the increase in an individual’s susceptibility to infection with strain *y*, when the antigenic distance to the infection memory *x* increases. Inspired by data for seasonal influenza, we assume a logistic, S-shaped increase for *K*_*s*_(*x − y*)

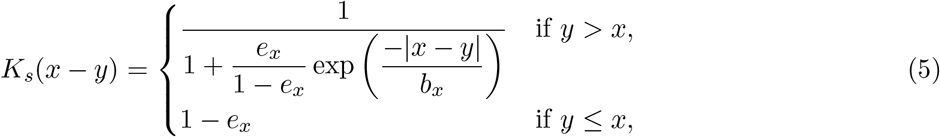

with a complete loss of cross-protection for distant strains, so that *K*_*s*_(*∞*) = 1 (Figure 2a, dashed line). The inverse slope near the inflection point, *b*_*x*_, describes the breadth of the protection against antigenically distinct strains, measured in antigenic units. The y-intercept, 0 *≤* (1 *− e*_*x*_) *≤* 1, describes the probability of reinfection with a strain that is antigenically identical to the infection memory, i.e., *y* = *x*. We assume the same probability for infection with an antigenically ‘older’ strain, *y < x*, to approximate protection against such older strains without storing all past infection memories (asymmetry in *x* in Figure 2a, dashed line).

**Figure 2.**
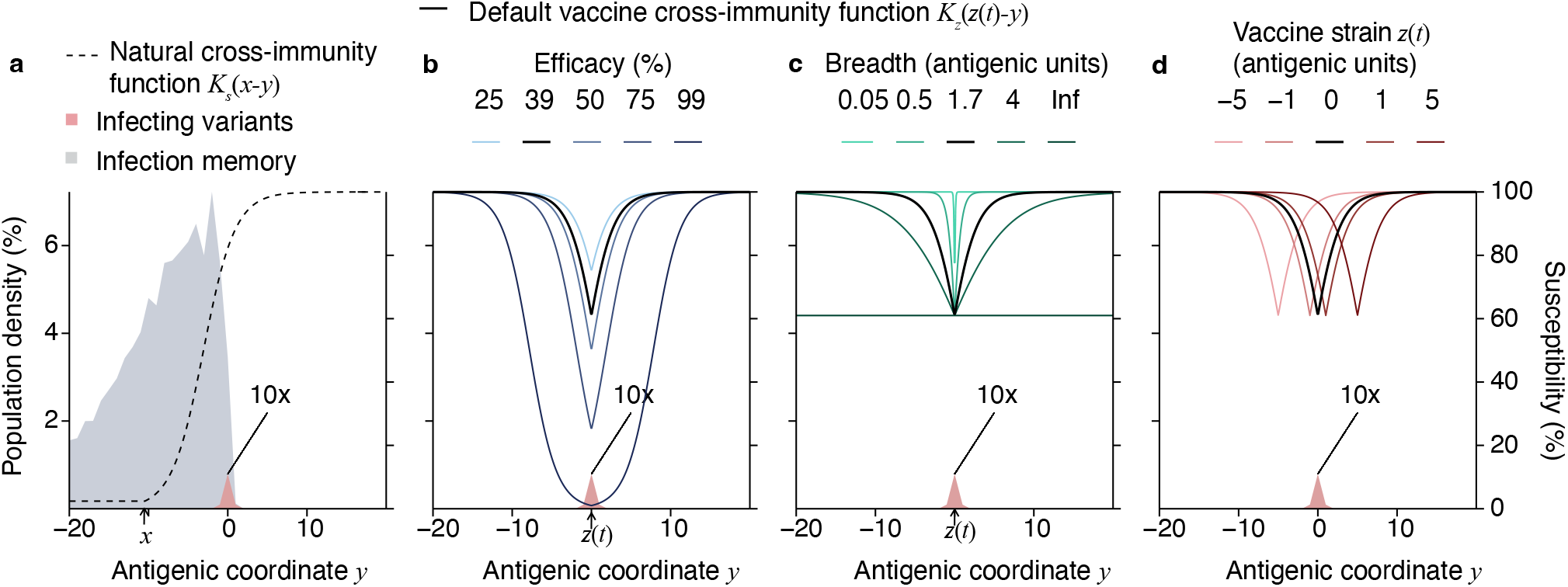
Cross-immunity functions. The cross-immunity functions describe the increase in susceptibility (right y-axis) when the antigenic coordinate of the infecting strain *y* (x-axis). The natural cross-immunity function, *K*_*s*_(*x − y*), (**a**, dashed line, right y-axis), compared to the effect of vaccine efficacy (**b**), breadth (**c**), and vaccine strain (**d**), in the vaccine cross-immunity function, *K*_*z*_(*z*(*t*) *− y*) (solid line, right y-axis). The cross-immunity functions can be compared to the distributions of the susceptible (grey, left y-axis) and infected individuals (red, left y-axis, 10 times enlarged for clarity), obtained under default parameter values without vaccination. The antigenic coordinate of the infecting strain *y* is measured relative to the mean of the infected distribution, which is set at *y* = 0. The natural cross-immunity is displayed for the average infection history of the susceptible population (*x* = *−*10.5 antigenic units). The black line indicates the default vaccine cross-immunity used (Table 1), and other colors indicate alternative scenarios tested.

In vaccinated individuals, the total susceptibility to any strain is obtained by multiplying the natural cross-immunity, *K*_*s*_(*x − y*), by an additional term for the vaccine-induced cross-immunity, *K*_*z*_(*z*(*t*) *− y*), assuming that natural infection memory and vaccine-induced immunity combine multiplicatively to reduce susceptibility

**Table 1.**
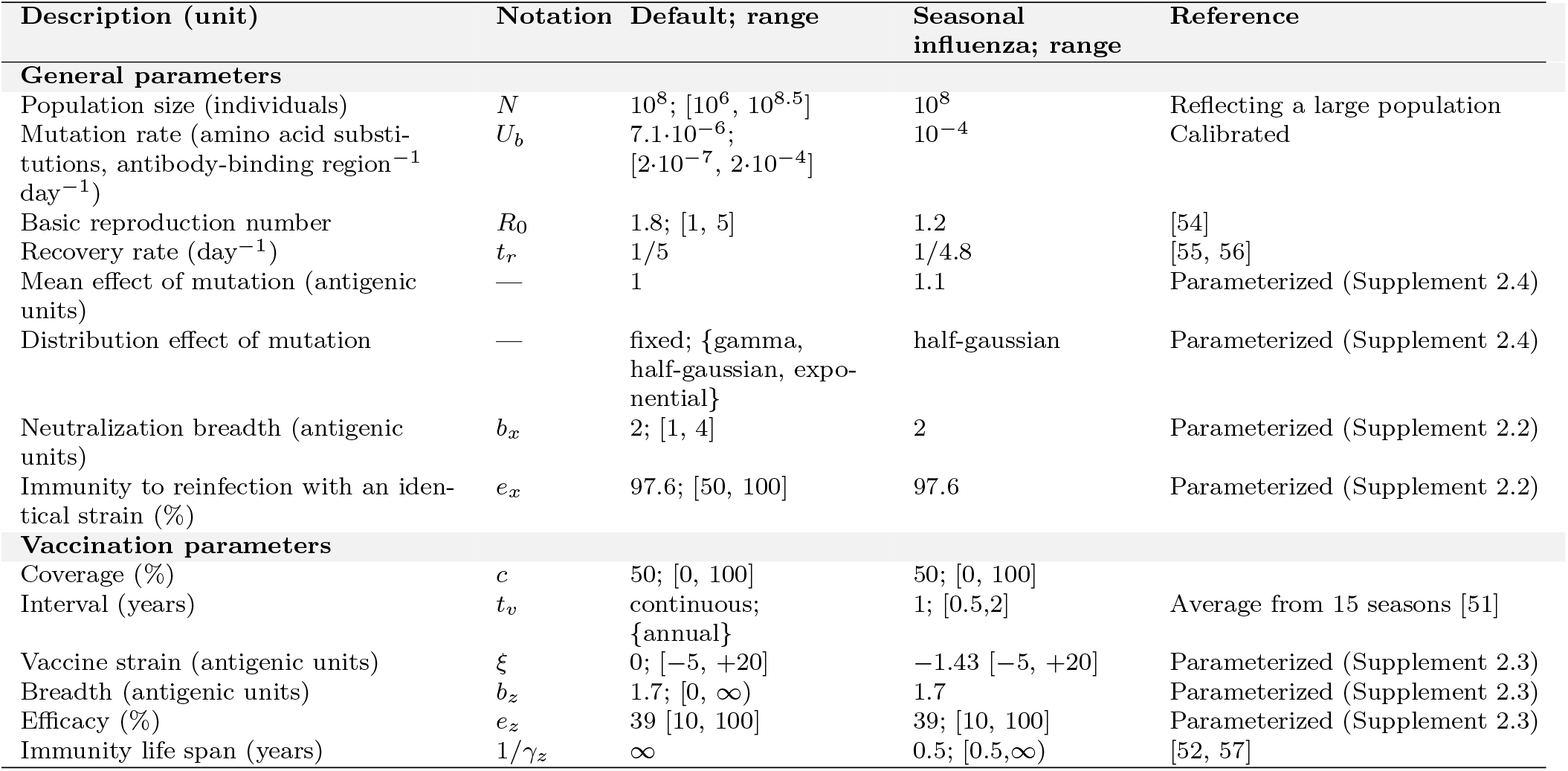
Model parameterization. For each parameter, the unit, notation, and values are specified for the default and seasonal influenza parameter sets. We also indicate the alternative parameter ranges explored in the main and sensitivity analyses, reflecting possible changes in vaccine programs.

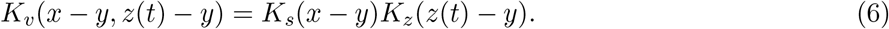

The vaccine cross-immunity function, *K*_*z*_(*z*(*t*)*−y*), describes the increase in susceptibility of vaccinated individuals with the increase in the antigenic distance between the infecting strain *y* and the vaccine strain *z*(*t*)

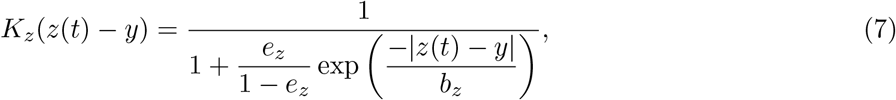

where 0 *≤ e*_*z*_ *≤* 1 is the vaccine efficacy, quantifying protection against infection with *z*(*t*), and *b*_*z*_ is the vaccine breadth, describing the breadth of vaccine protection against more distant strains (Figure 2b-d, solid lines). In contrast to natural cross-immunity, the vaccine cross-immunity decays for both ‘newer’ (*y > z*(*t*)) and ‘older’ (*y < z*(*t*)) strains, assuming that past vaccinations do not confer lasting immunity. For each vaccination event, a new vaccine strain *z*(*t*), i.e., the target of the vaccine, is selected at the coordinate

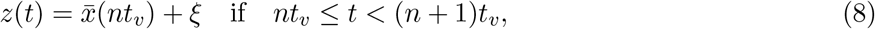

where *ξ* is the distance between the vaccine strain and the mean of the infected wave, 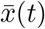, that is, the mean antigenic coordinate of all circulating strains

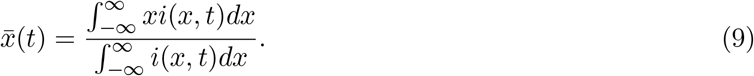

### 2.5. Data and parameterization

We used two parameter sets in our model simulations: a default set for the primary and sensitivity analyses, and a separate set for a seasonal influenza A/H3N2 case study (Table 1). For both parameter sets, we compared a no-vaccination scenario with multiple vaccination scenarios, varying vaccine-related parameters around their default values. Some parameters were estimated directly from data, whereas others were derived from the literature.

For the default parameter set, pathogen- and individual-specific parameters unrelated to vaccination were chosen to reflect the dynamics of a generic rapidly evolving pathogen [33, 43]. For simplicity, each mutation was assumed to advance the pathogen by a fixed step of one antigenic unit. Vaccination coverage was fixed at 50%, with the vaccine strain centered at the mean of the antigenic wave (*ξ* = 0) and no explicit waning of vaccine-induced immunity (*γ*_*z*_ = 0), as vaccine-induced immunity was updated at each time step. The parameters governing cross-immunity were set equal to those used in the seasonal influenza parameterization described below.

To inform the seasonal influenza parameter set, specifically the cross-immunity functions and the antigenic effects of mutations, multiple data sources were used. First, we used results from Hemagglutination Inhibition (HI) assays for 270 antisera and 15,804 influenza A/H3N2 strains collected by the World Health Organization Global Influenza Surveillance Network and published in [44], with data deposited on [45]. The outcome of this assay, the HI titre, measures the ability of antibodies in blood antiserum to neutralize influenza virus and is widely recognized as a correlate of protection. When measured across multiple antisera and strains, these titers can be used to calculate antigenic distances between strains, expressed in logarithmically scaled HI titre units (log_2_HI) [1]. Using the GenBank, we retrieved amino acid sequences of the hemagglutinin head domain, the region in which most antigenic mutations occur, for 1,378 strains [44, 46]. These data were used to quantify the distribution of antigenic effects associated with single amino acid mutations. On average, a single mutation produced an antigenic change of 1.3 log_2_HI units, with a right-skewed distribution (Figure S2).

To estimate the relationship between neutralization titre and susceptibility to infection, we used human and animal challenge studies in which HI titres were measured prior to exposure [47, 48]. Combining these results with the antigenic distance data, we found that immunity to homologous reinfection was high but not sterilizing (97%), and susceptibility increased with antigenic distance.

To parameterize vaccine-induced cross-immunity, we used data from vaccine challenge studies [49, 50] and longitudinal cohort studies, reporting both vaccine efficacy and antigenic distance between vaccine and infecting strains, systematically compiled in [51] for 15 calendar years. Vaccine efficacy against the vaccine strain was estimated at 39%, and vaccine breadth at 2.0 log_2_HI units. These data also indicated that vaccine strains lagged behind circulating strains by an average of 1.4 log_2_HI units. More details on calibration methods are provided in the Supplement 2.

Other vaccine-related parameters were chosen to reflect global influenza vaccination practices. Vaccination was assumed to occur annually [12], with vaccine-induced immunity waning over 6 months [52]. Lastly, we used a default vaccine coverage of 50%, exceeding the estimated global average of approximately 10% [53], in order to obtain more pronounced effects when varying other vaccine parameters. The basic reproduction number (*R*_0_ = 1.2) and mutation rate (*U*_*b*_ = 10^*−*4^ amino acid substitutions per antibody binding region per day) were calibrated to yield an annual infection incidence between 10 and 15%, consistent with global influenza estimates [44, 12].

### 2.6. Vaccination scenarios

We examined the impact of both implementation characteristics, linked to public health decisions, and biological characteristics, determined by vaccine design. For implementation, we varied vaccine coverage *c* and compared two deployment frequencies 1*/t*_*v*_: (i) continuous vaccination at each time step, which corresponds to one recovery period, obtained by substituting *δ*(*t − nt*_*v*_) = 1 in Equations 2 and 3; and (ii) annual vaccination, defined by *t*_*v*_ = 1 year.

For biological characteristics, we varied vaccine efficacy *e*_*z*_ from imperfect to fully sterilizing vaccines with 100% efficacy, and cross-immunity breadth *b*_*z*_ from strain-specific to broadly neutralizing vaccines, including a universal vaccine with infinite breadth. Finally, we varied the antigenic distance *ξ* between the vaccine strain *z*(*t*) and the mean position of the infected wave, 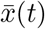 (Figure 2b-d). The parameter ranges explored for all vaccination scenarios are provided in Table 1.

### 2.7. Simulation outcomes

Simulations were initialized from pre-generated steady-state traveling waves, equilibrated under each parameter set to avoid transient effects (Supplement 1.1). In most cases, a single parameter was varied while others were held fixed at their default values. The main outcomes were annual infection incidence, defined as the proportion of the population infected per year, and the speed of antigenic evolution, defined as the displacement of the mean of the infected wave through the antigenic space, measured in antigenic units. A simulation was declared extinct when the total infection density dropped below the equivalent of one individual. Simulations ran for 40 years and were repeated 100 times to capture stochastic variability. Excluding simulations that went extinct, medians and interdecile ranges, defined as the 10th to 90th percentiles, are reported. Results are shown as absolute values and are compared to the no-vaccination scenario, unless stated otherwise.

### 2.8. Sensitivity analyses

To evaluate the robustness of our results and explore potential variations under alternative pathogen and modeling assumptions, we systematically varied: (i) general parameters specific to the pathogen and host population; (ii) simulation parameters, such as the time step and simulation duration, including analyses conducted within a single outbreak under varying initial susceptibility profiles; and (iii) the structure of the vaccination model.

For the latter, we evaluated a model with waning immunity, a model with multiple vaccination compartments, in which past vaccination events were retained, and a consistent vaccination scenario, in which vaccines were preferentially allocated to individuals previously vaccinated rather than assigned at random, as in the default model.

For each alternative parameter set and model structure, we compared the no-vaccination scenario with the default vaccination parameters. More details on the sensitivity analyses are provided in the Supplement 3.

### 2.9. Implementation

The model was implemented in the R programming language (Version 4.4.3). The antigenic space was discretized onto a one-dimensional lattice (Supplement 1.4). Equations 1-3 were solved numerically using Euler’s method. The time step, *t*_step_, was set to one recovery period *r* for the default simulations and to one day for the influenza case study.

## 3. Results

### 3.1. Traveling wave in the absence of vaccination

In the absence of vaccination, numerical simulations using default parameterization for a rapidly evolving pathogen showed that the distribution of infected individuals, stratified by the infecting strain, forms a narrow traveling wave in antigenic space (Figure 2a and Figure S3). Most circulating strains cluster within a limited antigenic range around a single dominant strain at any given time, with, on average, four strains accounting for about 95% of infections. The susceptible population, stratified by infection memory, trails the infected distribution at the same speed. Its distribution exhibits a sharp drop near the peak of the infected distribution, reflecting recovery of infected individuals, and a slowly decaying tail due to reinfection of susceptible individuals. The annual infection incidence was 9% (IQR: 8.9% - 9.4%), and the speed of antigenic evolution was 1.39 antigenic units per year (IQR: 1.36 - 1.45).

### 3.2. Impact of vaccine implementation characteristics

Next, we examined how these dynamics changed upon the introduction of vaccination under the default vaccination parameters. We considered two vaccination scenarios: continuous vaccination, i.e., after every recovery period, and annual vaccination (Figure 3). Under continuous vaccination, the shape of the traveling waves and the outbreak periodicity remained similar to the no-vaccination scenario (Figure S4a). However, increasing vaccination coverage led to a subtle decline in annual infection incidence, and at 100% coverage, all simulations went extinct. Interestingly, while infection incidence decreased, the speed of antigenic evolution, defined as the displacement of the mean of the infected wave through antigenic space over time, increased by up to one-third at 75% coverage. In contrast, annual vaccination caused more frequent extinctions, and for coverage levels of 50% or higher, no stable simulations could be established. This effect occurs because annual vaccination acts as a sudden perturbation to the system, particularly when deployed shortly after an outbreak, when the susceptible population is already low.

**Figure 3.**
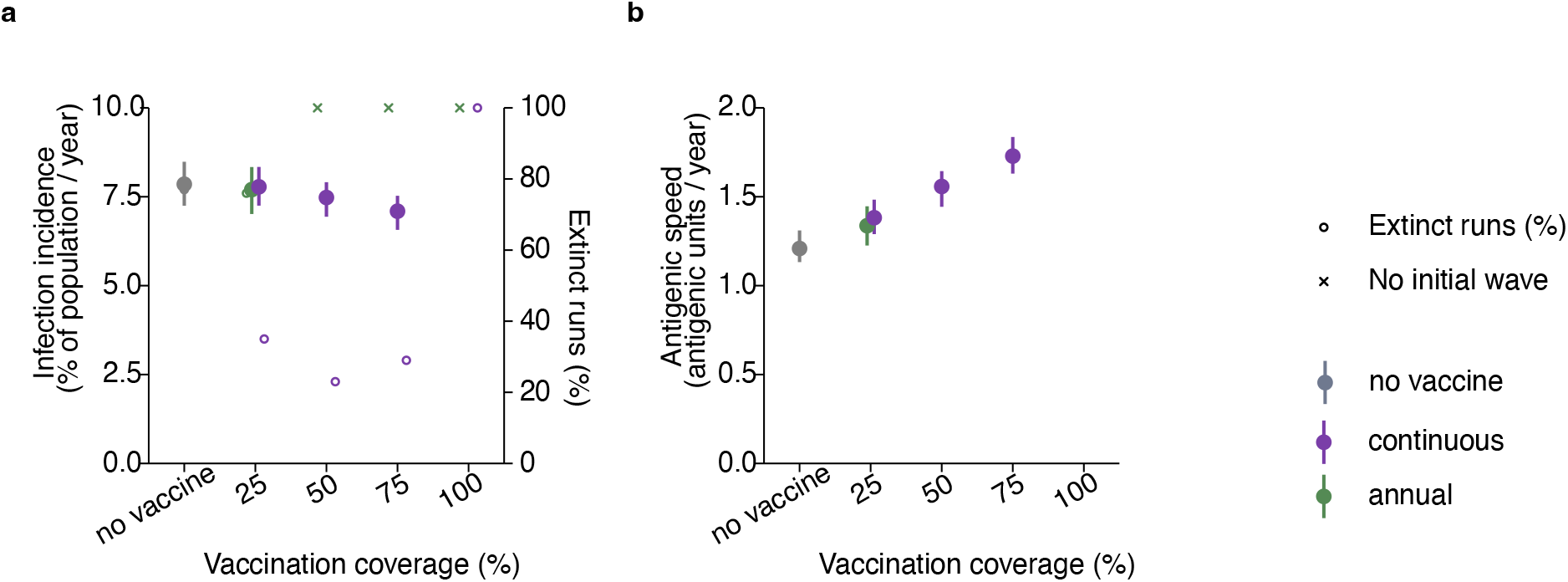
Impact of vaccine implementation characteristics. Impact of varying vaccination coverage and frequency on annual infection incidence (**a**, left y-axis) and the percentage of simulation runs resulting in pathogen extinction (**a**, right y-axis, open circles); crosses indicate cases where no stable traveling wave could be established. (**b**) The speed of antigenic evolution. Outcomes were averaged over a 40-year simulation period. Results are shown for continuous vaccination (purple), annual vaccination (green), and no vaccination (grey). Points represent the median, and the error bars represent the interdecile range across 100 stochastic simulation runs that did not go extinct. See Table 1 for parameter values.

### 3.3. Impact of biological vaccine characteristics

In addition to implementation characteristics, we examined how intrinsic biological properties of vaccines, such as efficacy, cross-immunity breadth, and vaccine strain, affect epidemiological and evolutionary dynamics. Each parameter was varied individually, while all other parameters were fixed at their default values.

First, we considered continuous deployment (purple dots in Figure 4). Increasing vaccine efficacy to 50% slightly reduced infection incidence, but the speed of antigenic evolution increased (Figure 4a,b). At an efficacy of 75% or higher, the pathogen went extinct. This pattern mirrors the effect of increasing vaccination coverage, as both higher efficacy and coverage linearly increase the population-level immunity per strain.

**Figure 4.**
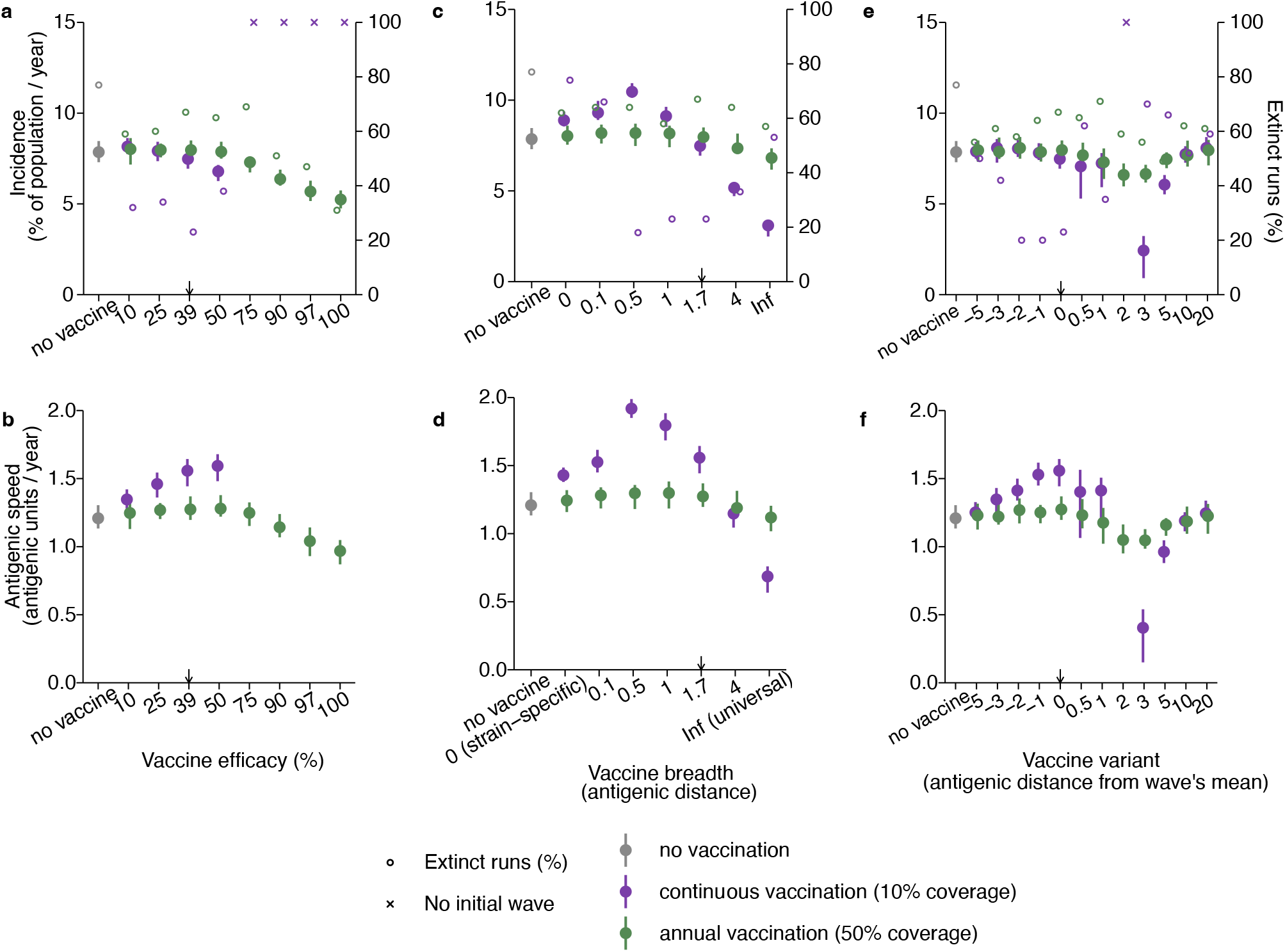
Impact of biological vaccine characteristics. Impact of varying vaccine efficacy, cross-immunity breadth, and vaccine strain on annual infection incidence (**a,c,e**, left y-axis) and the percentage of simulation runs resulting in pathogen extinction (right y-axis, open circles); crosses indicate cases where no stable traveling wave could be established. (**b,d,f**) The speed of evolution in antigenic units per year. Results are shown for continuous vaccination (purple), annual vaccination (green), and no vaccination (grey). Outcomes were averaged over 40 years. Points represent the median, and error bars represent the interdecile range across 100 stochastic simulation runs that did not go extinct. Small arrows indicate default vaccination parameter values (Table 1).

Next, we varied the cross-immunity breadth of the vaccine (Figure 4c,d). A strain-specific vaccine that protects against a single strain only, had little impact on infection incidence or evolutionary speed, because susceptibility remains high for most circulating strains. Increasing the breadth to 0.5 antigenic units accelerated antigenic evolution. A vaccine with such a narrow breadth protects against most circulating strains but offers little protection beyond that range, enhancing the growth advantage of emerging strains. Figure 2c shows that this cross-immunity function closely matches the width of the infected wave. Notably, in this scenario, a modest increase in incidence relative to no vaccination was observed. As the vaccine breadth increased beyond 0.5 antigenic units, both incidence and evolutionary speed decreased and eventually plateaued when approaching a universal vaccine that protects against all strains. In this scenario, the vaccine cross-immunity is flat (Figure 2c), such that new strains gain no fitness advantage by evading vaccine-induced immunity. Therefore, vaccination imposes no additional selection pressure for antigenic escape. The observed reduction in infection incidence of about 50% relative to no vaccination thus reflects the direct protective effect of vaccination, without confounding from accelerated antigenic evolution, highlighting how vaccine-driven evolution can partially offset vaccination benefits.

Finally, we varied the vaccine strain (Figure 4e,f). When the vaccine strain was selected at the mean of the infected wave, the acceleration in evolutionary speed was largest. Selecting the vaccine strain 2 antigenic units ahead of the wave’s mean prevented the establishment of a stable traveling wave. Vaccines targeting strains at the front of the wave lowered the fitness advantage of escape mutants, reducing both the speed of antigenic evolution and incidence, and frequently driving the pathogen extinct. Similar effects were observed for vaccines targeting strains 3 antigenic units ahead of the wave’s mean. For vaccines targeting strains further ahead, the vaccine-induced protection against circulating strains declined, and infection incidence and speed approached values without vaccination.

These analyses were repeated for annual vaccination at 10% coverage (instead of 50%) to avoid extinction. Lower coverage reduced the impact of vaccination, i.e., the pronounced increases in infection incidence and the speed of antigenic evolution were not observed, although the overall qualitative patterns remained similar.

### 3.4. Modeling vaccination to seasonal influenza A/H3N2

We further investigated the effect of vaccination in a case study of seasonal influenza A/H3N2, using parameters estimated from data (Supplement 2). Simulation results showed that influenza vaccines parameterized to global averages did not accelerate antigenic evolution compared to a no-vaccination scenario (Figure 5). However, high vaccination coverage and vaccine strains better matched to circulating strains could increase the speed of antigenic evolution by at most few percent. Overall, the effects of vaccine efficacy, breadth, and vaccine strain selection were consistent with those observed in the default scenario. For seasonal influenza, more frequent vaccination reduced the probability of pathogen extinction and amplified the overall impact on dynamics, leading to a stronger reduction in infections while slightly increasing the speed of antigenic evolution. Additionally, slower waning of vaccine-induced immunity slightly increased the impact of vaccination. Importantly, we observed no vaccine-induced increase in incidence.

**Figure 5.**
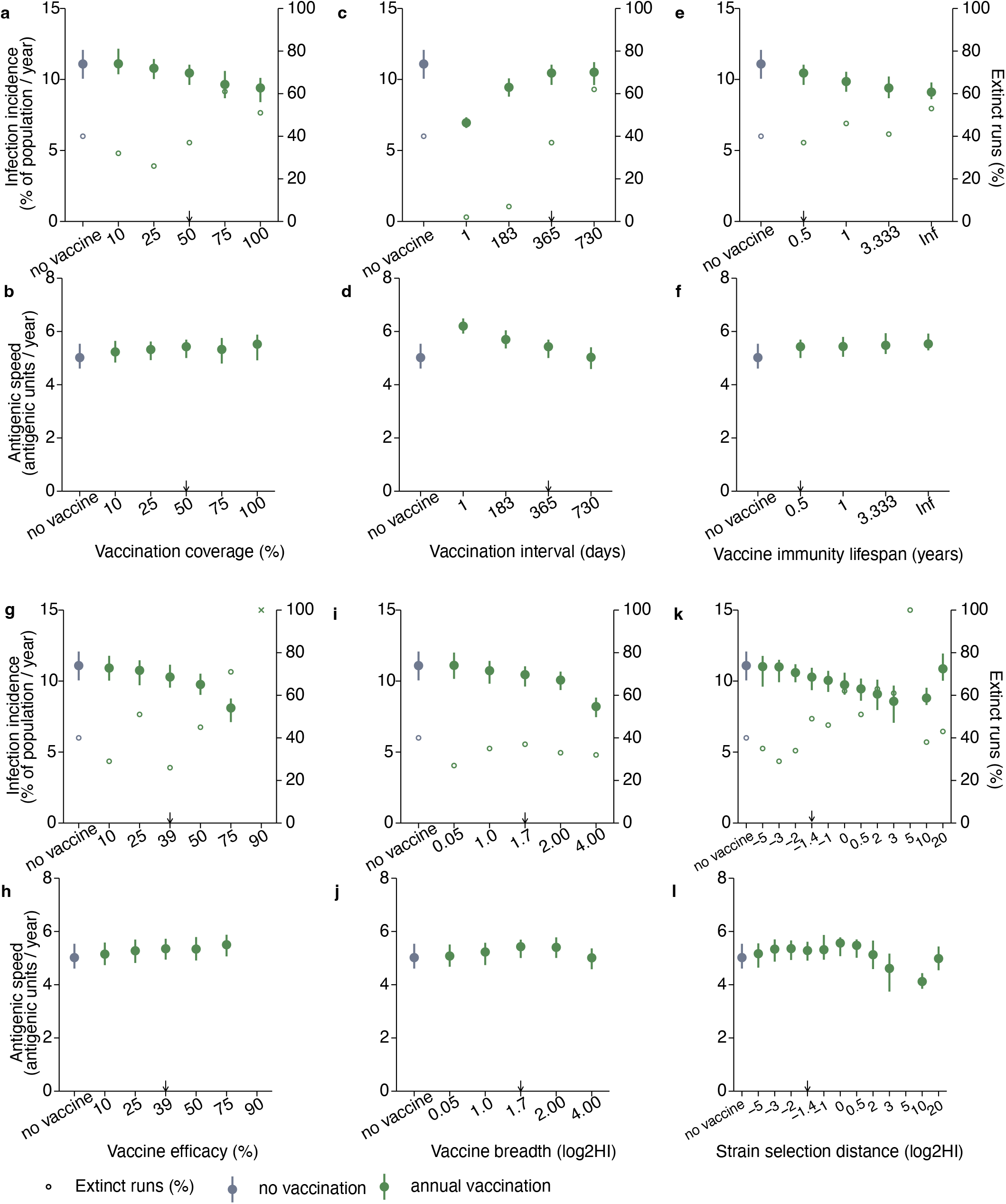
Impact of vaccine characteristics on seasonal influenza. Impact on annual infection incidence (**a,c,e,g,i,k**, left y-axis) and the percentage of simulation runs resulting in pathogen extinction (right y-axis, open circles); crosses indicate cases where no stable traveling wave could be established. Panels (**b,d,f,h,j,l**) show the speed of antigenic evolution. Outcomes were averaged over a 40-year simulation period and compared to the no-vaccination scenario (grey). Points represent the median, and error bars represent the interdecile range across 100 stochastic simulation runs that did not go extinct. See ‘seasonal influenza’ in Table 1 for parameter values.

### 3.5. Sensitivity to model assumptions and parameters

Model dynamics, even without vaccination, were sensitive to pathogen- and host-specific parameters (Figure S8). Larger population sizes, higher mutation rates, increased *R*_0_, and reduced efficacy or breadth of natural immunity all led to higher infection incidence and faster antigenic evolution. However, in none of these scenarios did vaccination substantially accelerate antigenic evolution further.

The antigenic effect of mutations was also influential. For distributions with greater variability, stable traveling waves did not emerge without vaccination, as large outbreaks of escape mutants were often followed by pathogen extinction (Figure S9). Vaccination mitigated these effects by increasing population immunity to escape mutants and thereby preventing both large outbreaks and subsequent pathogen extinction.

Simulation outcomes were robust to changes in key simulation parameters, such as time step size, antigenic coordinate length, and total simulation duration (Supplement 3.2 and Figure S6). We additionally explored the effect of vaccination in a single epidemic outbreak, using simulations over a one-year period, initiated with a single strain and varying profiles of population-level susceptibility (Supplement 3.3 and Figure S7). In the absence of vaccination, escape strains failed to establish during the outbreak. Interestingly, vaccination deployed at the start of the outbreak and targeted at the initial strain reduced the outbreak size but facilitated the establishment and further emergence of escape strains, particularly under conditions of narrow cross-immunity breadth and high coverage. Although this acceleration offset the reduction in incidence, it never increased incidence relative to the no-vaccination scenario.

Alternative models with multiple vaccination compartments, or with consistent rather than random vaccine allocation, did not produce a substantially greater increase in evolutionary speed or incidence than those observed in the baseline scenario (Figure S5). We also evaluated how dynamics altered when incorporating the waning of immunity. When immunity lasted for an individual’s lifetime, reflecting birth and death processes, dynamics were similar to the baseline. By contrast, for an immunity lifespan of 5 years, vaccination produced a greater acceleration of antigenic evolution, likely due to the increased selection pressure imposed by vaccination relative to natural immunity.

## 4. Discussion

Vaccination plays a key role in preventing transmission of rapidly evolving pathogens such as seasonal influenza and coronaviruses, particularly given recent advances in vaccine development and implementation programs. However, concerns have been raised regarding the potential for vaccination to accelerate antigenic evolution. In this study, we investigated how vaccination influences both the speed of antigenic evolution and infection incidence for rapidly evolving pathogens. Using a stochastic, multi-strain, population-level compartmental model, we systematically evaluated the impact of vaccine implementation characteristics (coverage and frequency) and biological properties (efficacy, breadth, and vaccine strain selection) on incidence and evolutionary speed.

In the absence of vaccination, the distribution of infected individuals formed a narrow wave traveling in antigenic space, driven by mutation and immune selection, consistent with multi-locus evolutionary theory [35, 58, 59, 60, 61, 62]. Introducing vaccination with high efficacy, coverage, or breadth generally reduced infection incidence and, in many cases, slowed antigenic evolution, consistent with previous studies [28, 19, 30, 17]. At sufficiently high coverage and efficacy, vaccination drove the pathogen to extinction. Such abrupt declines in incidence have previously been termed the “drift cliff” [19, 17]. This phenomenon can be explained by a positive feedback mechanism: reduced incidence lowers the chance of emergence of novel strains, which further suppresses transmission. Somewhat counterintuitively, less frequent (annual) vaccination led to a larger reduction in infections than continuous vaccination. Infrequent vaccination acted as a perturbation to the system, increasing the probability of pathogen extinction. However, such abrupt extinctions may be less likely in real-world settings, where vaccination is implemented more gradually. Vaccines targeting strains antigenically ahead of the traveling wave were also more likely to reduce transmission and drive pathogen extinction. In reality, however, for pathogens evolving in multidimensional antigenic space, incorporating all potentially emerging strains into a single vaccine may not be feasible.

When vaccination allowed pathogen persistence due to low coverage or low efficacy and conferred a growth advantage to escape strains, we observed a modest acceleration of antigenic evolution. Consistent with this finding, the two-strain SIR model by Rella et al. [31] showed that vaccination at coverage levels insufficient to interrupt transmission favored the emergence and establishment of vaccine-resistant strains. The largest relative increase in antigenic evolution in our model reached up to one-third of the no-vaccination baseline and occurred when the vaccine strain matched the mean of the infected wave while vaccine breadth was narrow. Such vaccines effectively protected against most circulating strains but provided little cross-protection beyond that range, thereby optimizing the selective growth advantage of newly emerging strains. Similarly, using a multi-strain SIR model in two-dimensional antigenic space with gamma-distributed mutational steps and a linear cross-immunity function, Wen et al. [19] found that vaccines with low to intermediate breadth could accelerate antigenic evolution. In contrast, vaccines with broader protection slowed antigenic evolution. Using a fitness model for SARS-CoV-2 informed by neutralization data, Meijers et al. [29] likewise showed that broader vaccines reduced evolutionary speed. In the limiting case of a universal vaccine, protection does not discriminate between strains, resulting in the largest reduction in incidence. Similarly, Arinaminpathy et al. [32] demonstrated in a multi-strain SIR model that continuous deployment of broadly protective vaccines could slow antigenic evolution in seasonal influenza. Our comparison of universal and narrower vaccines suggests that vaccine-induced evolution can partially offset reductions in infection incidence. Without vaccine-induced selective pressure, the impact of vaccination would likely be considerably greater.

Although we explored a broad range of vaccine and pathogen parameters, we cannot exclude the possibility that the true maximum long-term increase in evolutionary speed or incidence under vaccine pressure may be higher than observed here. In parallel work, our baseline model with continuous vaccination was analyzed analytically [43]. An important difference is that, in certain parameter regimes (e.g., high coverage), analytical results suggest a larger vaccine-induced increase in evolution and incidence, whereas in our stochastic simulations these regimes more often led to extinctions.

It is also important to consider short-term antigenic evolution, particularly in scenarios where our long-term simulations result in permanent extinctions. For many pathogens, transmission may cease temporarily due to epidemiological or ecological factors. When a novel strain is reintroduced from other regions or reservoirs [56], the probability of a subsequent outbreak strongly depends on the remaining cross-immunity in the population. In our sensitivity analyses of such single outbreaks, we observed a shortterm acceleration of evolution under certain vaccination scenarios, depending on pre-existing population immunity. Although this acceleration offset the reduction in incidence, it never increased incidence relative to the no-vaccination scenario. These findings are consistent with the increased establishment probability of vaccine escape strains reported by Gandon et al. [30] in a two-strain model for a pandemic setting.

We further performed a case study of seasonal influenza A/H3N2, using parameters informed by challenge studies, neutralization assays, and sequence data. Our results suggest that current influenza vaccines, which generally have low efficacy and limited coverage worldwide [51, 53], exert minimal impact on long-term incidence and antigenic evolution. When vaccine strains were better matched to circulating strains, we observed a modest acceleration of antigenic evolution, but no increase in incidence. These results indicate that ongoing efforts to improve vaccine efficacy or strain selection, including enhanced strain prediction tools, expanded surveillance, improved delivery methods, higher antigen doses, adjuvants, and next-generation vaccine platforms [63] should be evaluated in light of their evolutionary consequences. In contrast, the development of vaccines with broader protection, such as multivalent formulations or vaccines targeting conserved epitopes, is more likely to reduce antigenic evolution [11].

Our findings were robust to variation in simulation parameters and alternative models of vaccination. Nonetheless, it must be noted that antigenic evolution and transmission are inherently stochastic processes, as reflected in the high variability in model outcomes. This variability highlights the challenges of predicting vaccine-driven evolutionary dynamics in real-world settings.

Our model has several limitations. First, it does not account for fitness costs of immune escape mutations. Including such costs would likely reduce the overall fitness advantage of escape strains and may therefore attenuate vaccine-induced acceleration of evolution suggested by our model [17]. Second, we focus on vaccines that reduce transmission and incidence rather than severe disease outcomes, and thus do not evaluate the evolution of virulence [64, 65]. However, the risk of virulence-increasing mutations may further heighten concerns regarding vaccine-driven evolution. Third, population heterogeneity, including age-related immune differences and contact patterns, is not considered, although such factors may affect the fitness advantage of escape strains, especially under targeted vaccination strategies [66, 67, 30]. Fourth, we assume an infinite one-dimensional antigenic space. Previous work has shown that influenza strains spontaneously form a one-dimensional evolutionary trajectory in multi-dimensional antigenic space [33, 56]. Results from our model are qualitatively similar to those from higher-dimensional frameworks [19]. However, other parameter regimes in higher-dimensional antigenic spaces may lead to qualitatively different dynamics such as speciation [68]. Fifth, the model omits seasonality and spatial spread, both of which are important for pathogens like seasonal influenza [41]. Finally, the effect of vaccination is sensitive to pathogen-specific model parameters. Our findings should therefore not be interpreted as arguments against vaccination, which remains a critical intervention for infectious disease control. Rather, to allow more precise evaluation of vaccine effects, the model should be extended to incorporate additional biological and epidemiological processes such as seasonality, mutational fitness costs, original antigenic sin, back-boosting, antibody-dependent enhancement, individual heterogeneity, and antigenic shifts [67, 17, 41].

Substantial progress has been made in modeling the epidemiological impact and cost-effectiveness of novel vaccines, for example, for influenza [14]. Future work should integrate vaccine-induced evolutionary dynamics into such evaluations, as our results indicate that accelerated antigenic evolution may partially offset vaccine-driven reductions in infection burden. In particular, when population-level vaccine effectiveness is unexpectedly low, pathogen-specific multi-strain models may help understand whether accelerated evolution contributes to reduced vaccine effectiveness.

In conclusion, our model shows that vaccination can effectively reduce both infection incidence and the speed of antigenic evolution in many scenarios. However, when vaccination permits sufficient transmission while conferring a selective fitness advantage to escape strains, it may accelerate antigenic evolution and, thereby, partially offset reductions in incidence. The potential for vaccine-driven evolution therefore warrants careful consideration in the design and deployment of new vaccines.

## 4.1. Acknowledgements

Funding: This work was supported by the NCOH Pandemic Preparedness Kickstarter (ZonMW Grant, #10710022210003). The present work originated in numerous discussions with Igor Rouzine. We are grateful to him for sharing his manuscript prior to publication.

## 4.2. Author contributions

G.R. supervised the study. M.S.W and G.R. developed the mathematical model and the scenarios. M.S.W carried out data analysis, estimated parameters, wrote computer code, conducted and planned simulations, and prepared figures. M.S.W. and G.R. wrote the manuscript.

## 4.4. Data availability

The model code, figure reproduction scripts, and associated data will be openly available at https://github.com/MyrtheW/vaccination_and_antigenic_evolution upon publication.

