## Supplement for "Impact of vaccination on the speed of antigenic evolution"

### Supplementary Material

Myrthe S Willemsen[1a,b], Ganna Rozhnova[a,b,c,d]

[a] *Julius Center for Health Sciences and Primary Care, University Medical Center Utrecht, Utrecht University, Utrecht, The Netherlands*
[b] *Center for Complex Systems Studies (CCSS), Utrecht University, Utrecht, The Netherlands*
[c] *BioISI—Biosystems & Integrative Sciences Institute, Faculty of Sciences, University of Lisbon, Lisbon, Portugal*
[d] *Faculty of Sciences, University of Lisbon, Lisbon, Portugal*

---

[1] Corresponding author:
Myrthe S Willemsen
Julius Center for Health Sciences and Primary Care
University Medical Center Utrecht
P.O. Box 85500 Utrecht
The Netherlands


### 1. Supplementary Methods

#### 1.1. Initial conditions and transient period

Here, we describe how we initialize the distributions of susceptible unvaccinated, susceptible vaccinated, and currently infected individuals, further referred to as 'initial waves'. The method described aims to avoid transient effects such as fluctuations that arise because initial conditions are far from the steady state, and extinction artifacts when initial waves result in an extinction event with a high probability shortly after initialization. For each parameter set, the following procedure was used to obtain the initial waves:

1. To start the simulation, select another initial wave from either the default parameter set or, if parameters are incrementally changed, select a previously computed wave from the most similar parameter set.
2. Simulate for 30 years with the parameter set of interest.
3. If the pathogen did not go extinct, record snapshots of the wave at years 10, 11, through 19. In case vaccination is periodic, record these immediately before vaccination events. This ensures that the forced timing of vaccination in the main simulations aligns with that of the steady state. The first 10 years of the simulation are excluded to allow the system to equilibrate under the new parameters, removing transient effects. The last 10 years are also excluded to exclude the possibility that the dynamics approach extinction.
4. If the pathogen did go extinct, repeat steps 1–3 up to 10 times. If available, use a different initial wave from the closest parameter set each time.
5. For each parameter set, use the 10 resulting initial waves as starting points for different runs of the main simulations.

#### 1.2. Genetic drift

Genetic drift describes the change in the frequency of a strain due to random events [1, 2], which is particularly important when the number of infections is small. To capture this effect, we track absolute numbers of infected individuals. When the number of recovered or newly infected inidivuals for a given antigenic coordinate falls below one hundred, it is replaced by a random draw from a Poisson distribution. Afterwards, we rescale the susceptible and vaccinated distributions proportionally to keep the population size $N$ constant.

#### 1.3. Mutations

The number of mutants generated by each strain is drawn from a Poisson distribution, where the expectation of the number of mutations is determined by the mutation rate $U_b$:

$$\text{rPois}\Big(\lambda = U_b i(x,t) N\Big)/N$$

For each mutant, the antigenic effect of the mutation $\Delta x$ is fixed at one or drawn from a probability distribution $P(\Delta x)$, which we vary between gamma, half-gaussian, and exponential. These alternative 'stepsize distributions' all had a mean of 1 to make the overall effect of mutations comparable to the fixed stepsize.

#### 1.4. Discretization of antigenic space

For computational tractability, the one-dimensional antigenic axis was discretized into a lattice. This lattice comprised of 200 bins of 1 antigenic unit each in the baseline model or 2000 bins of 0.1 antigenic units for the alternative stepsize distributions, including the seasonal influenza case study, and the integrals in Equations 1 - 4 in the main text were approximated with sums. The first bins of the susceptible unvaccinated and vaccinated compartments represent individuals with no infection memory $s(x = -\infty)$,

$v(x = -\infty)$. Whenever the infected wave reached the end of the lattice, we shifted all three waves backward along the lattice by 10% of the total length of the lattice. Sometimes, susceptible unvaccinated and vaccinated individuals with old infection memories in the tail of the waves would not fit on the lattice after shifting.We then redistributed these individuals between the coordinates representing no infection memory and the oldest strain, in a way that ensured that the total force of infection remained unchanged.

#### 2. Parameterization of seasonal influenza

Here, we describe how we used the data in Section 2.5 in the main text to parameterize cross-immunity functions and distributions of the antigenic effects of mutations for seasonal influenza A/H3N2. We first describe how we correlated inhibition titers with infection probability, then how we parameterized the natural cross-immunity, the vaccine cross-immunity, and the stepsize distribution.

##### *2.1. Hemagglutination Inhibition titers and the probability of infection*

Hemagglutination Inhibition (HI) titer values approximate the capacity of an antiserum to neutralize the influenza virus. While not flawless, HI titers are the standard correlate of protection and are widely used to compare the antigenic similarity of influenza strains. As a first step in characterizing the cross-immunity functions, we correlated HI titers with the probability of infection, using data from challenge studies [3, 4, 5]. These studies first measured the HI titers for a certain strain, then challenged these individuals with the same strain, and reported the number of cases for certain intervals of pre-challenge HI titers. We combined data from these studies and used iteratively reweighted least squares to fit a logistic regression curve (midpoint: 2.62, growth rate: $-0.49$) to the binary infection outcomes and the logarithmically scaled HI titer ($\log_2$HI) at the midpoint of each interval.

##### *2.2. Natural cross-immunity function*

In order to convert the relationship between HI titer and the probability of infection into a cross-immunity function, we needed information on HI titers against a strain of past infection. Using the GISN data set and regular expressions, we could match 698 strain names with antisera annotations, which denote the strain of immunization. On average, these pairs had a HI titer of 9.91 (95% CI $9.82 - 9.99$). We subtracted this value from the $\log_2$HI titers described in Section S2.1, to obtain a natural cross-immunity function with a breadth of $b_x$ of 2.0 $\log_2$HI and immunity to reinfection with an identical strain $e_x$ of 97.6% (Figure S2). This aligns with earlier findings that immunity against seasonal influenza is non-sterilizing [6, 7].

##### *2.3. Vaccine cross-immunity function*

We assumed that vaccine cross-immunity multiplicatively combines with natural immunity to reduce susceptibility. To quantify this relative risk reduction, we pulled different data sources. For all studies, we collected measures of HI titer and either infection probability or efficacy. First, we used data from an equine challenge study. Previously unexposed horses were vaccinated and then challenged with an antigenically different equine influenza strain [8]. Second, we used data from human challenge studies, reviewed by Basta et al. [9]. These studies vaccinated individuals against influenza A/H3N2 and then challenged them with the vaccine strain. Third, we used data from human observational studies [10, 11]. These studies reported vaccine efficacy and the antigenic distance between the circulating and vaccine strains.

For vaccine-induced cross-immunity, we assumed a logistic function fitted to pooled data. Unlike natural cross-immunity, vaccine-induced protection was assumed to be symmetric, with equal decay against strains antigenically ahead of or behind the vaccine strain.

##### *2.4. Antigenic effects of mutations in seasonal influenza*

Random amino acid substitutions in viral epitopes can reduce the ability of existing antibodies to neutralize mutant strains. This occurs to varying degrees depending on the change in charge, size, hydrophobicity, and conformational change. The variable effect sizes of mutations have previously been modeled using various theoretical distributions [12, 13, 14] and have been shown to influence dynamics [15, 16]. Here, we characterized the effect of single amino acid mutations on the antigenic distance for seasonal influenza, using the GISN data set and amino acid sequences from GenBank. For 10,960 pairs of strains that differed by exactly a single amino acid substitution, we calculated the antigenic distance as the $\log_2$HI differences averaged among sera. We fitted a half-Gaussian distribution with mean 1.3, which fit the data better than exponential or gamma distributions (Figure S2).

#### 3. Sensitivity analyses

To assess the robustness of our findings and explore the potential for different outcomes under alternative pathogen and modeling assumptions, we systematically varied key parameters and model structures. Here we discuss methods and results for: (i) general parameters (Section 3.1); (ii) simulation parameters (Sections 3.2 and 3.3); and (iii) the structure of the vaccination model (Sections 3.4, 3.6 and 3.5).

##### *3.1. Sensitivity to general parameters*

To assess how key baseline parameters, which may differ between pathogens or host populations, influence model outcomes, we performed a one-way sensitivity analysis (Figure S8). We evaluated the effect of the mutation rate, basic reproduction number ($R_0$), susceptibility to reinfection, population turnover, and the breadth of natural immunity on both infection incidence and the speed of antigenic evolution, up to the point where the alternative parameter value resulted in pathogen extinction. The model dynamics were sensitive to the average mutation rate and basic reproduction number: lower values led to pathogen extinction, while higher values increased both incidence and evolutionary speed, and reduced the impact of stochasticity. For neither of these scenarios, the effect of vaccines is altered relative to the scenario without vaccines.

When protection against reinfection was reduced, reinfections became more common, leading to higher incidence and making sustained antigenic evolution less critical for pathogen persistence. In such a scenario, in which natural immunity is reduced, the relative contribution of vaccination to population-level immunity increases. Consequently, vaccination showed a greater relative impact in reducing incidence, though the effect on evolutionary speed is less pronounced. Similarly, when the breadth of natural cross-immunity was narrower than that conferred by vaccination, vaccination resulted in a larger reduction in incidence. For large breadths we did not find stable initial waves.

Finally, we considered the effect of the stepsize distributions, which determine how much a single mutation alters the antigenic distance and allows immune escape. In the baseline model, we used a fixed stepsize distribution for antigenic mutations, where each mutation resulted in a change of one antigenic coordinate. To explore the impact of different stepsize distributions on evolutionary dynamics, we also considered alternative distributions. These alternative distributions all had a mean of 1 to make the overall effect of mutations comparable to the fixed stepsize, including gamma distributions with variances 0.1, 0.5 and 1 (exponential), and a half-Gaussian distribution. Figure S9 shows that for stepsize distribution with larger variability (half-gaussian and gamma with variances 0.5 and 1), no stable simulations were obtained in the absence of vaccination. These stepsize distributions facilitate the emergence of escape mutants for which population-level immunity is so low that large outbreaks occur. Following these outbreaks, immunity is so high that the pathogen goes extinct. Vaccination, however, when deployed at every time step, rapidly follows escape strains and provides population-level immunity that can prevent these large outbreaks and extinctions from occurring. It remains notable how the incidence and speed of antigenic evolution increase with greater variability in the stepsize distribution.

##### 3.2. Sensitivity to simulation parameters

We evaluated the sensitivity of our results to key simulation parameters, including the time step, the total duration of the simulation, and the length of the antigenic coordinate (Figure S6). Reducing the time step of the Euler method (default: one recovery period) led to a slight increase in infection incidence and a corresponding decrease in the frequency of extinction events, as well as a modest increase in evolutionary speed; however, overall differences between vaccination scenarios did not change. Moreover, simulation outcomes were robust to variations in the length of the antigenic coordinate and the total simulation time. Extending the simulation time, as expected, increased the probability of pathogen extinction due to the cumulative effect of stochastic events.

##### 3.3. Short-term evolution

To investigate the short-term impact of vaccination, we simulated outbreaks starting from various initial susceptibility profiles, varying both the shape of the distribution and the overall population-level immunity to the initial infecting strain. For each parameter combination, we performed 10 simulation runs over a maximum of one year and averaged the results across all runs, including those where the pathogen went extinct. For each scenario, we calculated the cumulative proportion of individuals infected during the outbreak (i.e., the annual infection incidence) and the maximum displacement of the mean antigenic coordinate of the infecting wave. Vaccination was targeted at the initial infecting strain and only performed at the start of the simulations. Baseline parameters in Table 1 in the main text were used. Next, we describe the initial conditions used and then the results.

Initial conditions were derived from the average steady-state susceptible and infected wave from simulations without vaccination in the baseline model (Figure S3). We fitted a gamma distribution to the susceptible wave (shape 3.12, scale 4.6) and additionally created a 'wide' distribution (scale five times larger) and a 'peaked' distribution (scale five times smaller). The initial number of infections was set to match the steady-state prevalence (0.11%), but all infections were concentrated at a single antigenic coordinate. We varied the antigenic coordinate of the infecting strain while maintaining constant population-level cross-immunity to the new strain. To ensure outbreaks would occur within the simulation time frame, simulations were performed with 1.25-, 1.5-, and 1.75-fold reductions in population-level immunity to the initial strain, compared to the average population-level immunity to all circulating strains in the main simulations. Collectively, the different initial waves represent a range of outbreak scenarios, reflecting populations with varying heterogeneity and magnitudes of pre-existing immunity (Figure S7).

First, we consider what happens during a single epidemic outbreak without vaccination. While a few new strains emerged, they did not establish. Intuitively, the outbreak size increased when the prior population-level immunity to the initial strain was reduced. The population with more heterogeneous immune memories had a slightly lower outbreak size.

Vaccination at the start of the outbreak, targeted at the initial strain, decreased the outbreak size and increased the speed progressively with larger vaccination coverage. The increase in speed was greater when the prior immunity was lower and less heterogeneous. The speed increase of 1.5 antigenic units was small in absolute terms and large in relative terms. When the vaccine was only effective for the initial strain without providing cross-immunity, the increase in speed was highest. When, instead, it also provided cross-immunity against escape mutants, the speed decreased. When the vaccine protected against all circulating strains, the incidence and speed both reduced.

##### 3.4. Waning of immunity

We considered the effect of the lifespan of immunity, due to immunity waning and population turnover. At rate $\gamma$, individuals in all compartments were moved to a fully susceptible state $s(x=-\infty)$:

$$
\begin{aligned}
\frac{\partial i(x,t)}{\partial t} &= -(r+\gamma)\, i(x,t) \\
&\quad + \beta\, i(x,t) \int_{y=-\infty}^{\infty} K_s(y-x)s(y,t) + K_v(y-x, z(t)-x))v(y,t)dy, \\
\frac{\partial s(x,t)}{\partial t} &= r\, i(x,t) - \beta s(x,t) \int_{y=-\infty}^{\infty} K_s(x-y)i(y,t)dy \\
&\quad + \gamma(\delta(x+\infty) - s(x,t)) + \gamma_z v(x,t) + \delta(t-nt_v)\left(v(x,t) - c[s(x,t)+v(x,t)]\right), \\
\frac{\partial v(x,t)}{\partial t} &= -\beta v(x,t) \int_{y=-\infty}^{\infty} K_v(x-y, z(t)-y)i(y,t)dy - (\gamma_z+\gamma)v(x,t) \\
&\quad + \delta(t-nt_v)\left(-v(x,t) + c[s(x,t)+v(x,t)]\right),
\end{aligned}
$$

where

$$\beta = R_0(r+\gamma)$$

Short immunity lifespans increased the infection incidence, because more fully susceptible individuals entered the population, allowing the pathogen to persist without requiring a high speed of antigenic evolution. In these scenarios, the effect of vaccination was diminished due to the rapid depletion of vaccinated individuals. However, for an intermediate immunity lifespan of 5 years the acceleration in evolution was at a maximum.

##### 3.5. Vaccination compartments

For computational efficiency, the baseline model considered the effect of a single vaccination event at a time, neglecting earlier vaccinations. If prior vaccinations still contribute to immunity, this approach may overestimate the fitness of antigenically older strains. To evaluate the effect of this assumption, we evaluated an alternative model, with $N = 2$, 3, or 4 vaccine compartments, tracking individuals vaccinated in any of the last $n$ rounds by their most recent vaccine strain. Here, $j$ indexes the vaccine compartments ($j = 1, \ldots, N$):

$$\frac{\partial s(x,t)}{\partial t} = r\,i(x,t) - \beta s(x,t) \int_{y=-\infty}^{\infty} K_s(x-y) i(y,t) dy$$
$$+ \delta(t - nt_v)(v_n(x,t) - c[s(x,t) + v_n(x,t)])$$
$$\frac{\partial v_1(x,t)}{\partial t} = -\beta v_1(x,t) \int_{y=-\infty}^{\infty} K_v(x-y, z_1(t)-y) i(y,t) dy$$
$$+ \delta(t - nt_v)(-v_1(x,t) + c[s(x,t) + v_n(x,t)])$$
$$\frac{\partial v_2(x,t)}{\partial t} = -\beta v_2(x,t) \int_{y=-\infty}^{\infty} K_v(x-y, z_2(t)-y) i(y,t) dy$$
$$+ \delta(t - nt_v)(-v_2(x,t) + v_1(x,t))$$
$$\frac{\partial v_N(x,t)}{\partial t} = -\beta v_N(x,t) \int_{y=-\infty}^{\infty} K_v(x-y, z_N(t)-y) i(y,t) dy$$
$$+ \delta(t - nt_v)(-v_N(x,t) + v_{N-1}(x,t))$$
$$\frac{\partial i(x,t)}{\partial t} = -r\,i(x,t)$$
$$+ \beta\, i(x,t) \int_{y=-\infty}^{\infty} K_s(y-x) s(y,t) + \sum_{j=1}^{N} (K_v(y-x, z_j(t)-x) v_j(y,t)) dy$$
$$\int_{-\infty}^{\infty} i(x,t) + s(x,t) + \sum_{j}^{N} v_j(x,t) dx = 1$$

Figure S5 shows that additional vaccination compartments have a similar effect to increasing vaccine coverage and efficacy. The dominant effect of additional vaccination compartments is that the overall number of individuals with a vaccine status is higher, explaining these patterns.

##### *3.6. Consistent vaccination*

In the baseline model, vaccination was randomly allocated at each vaccination event ('random vaccination'). Here, instead, it is consistently allocated to individuals remaining in the vaccination compartment, that is, those vaccinated at the previous vaccination event who did not become infected in the meantime ('consistent vaccination'). To maintain the desired overall coverage $c$ at each vaccination round, we supplement the vaccinated compartment by adding $c'$ newly vaccinated individuals, accounting for those who left the vaccinated compartment due to infection or waning:

$$\frac{\partial s(x,t)}{\partial t} = r\,i(x,t) - \beta s(x,t) \int_{y=-\infty}^{\infty} K_s(x-y) i(y,t) dy$$
$$+ \gamma_z v(x,t) - \delta(t - nt_v) c' s(x,t)$$
$$\frac{\partial v(x,t)}{\partial t} = -\beta v(x,t) \int_{y=-\infty}^{\infty} K_v(x-y, z(t)-y) i(y,t) dy - \gamma_z v(x,t)$$
$$+ \delta(t - nt_v) c' s(x,t)$$

where

$$c' = \frac{\int_{x=-\infty}^{\infty} c[s(x,t) + v(x,t)] - v(x,t) dx}{\int_{x=-\infty}^{\infty} s(x,t) dx}$$

Compared to random vaccination, the waves of infected and susceptible individuals differed as, individuals who are repeatedly vaccinated experience fewer infections and accumulate distinct immune memories compared to those who are never vaccinated. Figure S5a shows a modest increase in the acceleration of antigenic evolution and no substantial change in annual infection incidence.

#### 4. Supplementary Figures

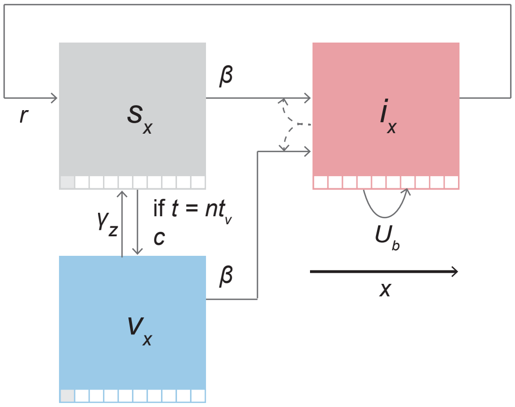


Figure S1: **Schematic of the multi-strain transmission model**. Individuals are classified as susceptible ($s_x$), infected ($i_x$), or vaccinated ($v_x$), each stratified by their most recent infection ($x$). Arrows indicate transitions: infection ($\beta$), recovery ($r$), vaccination ($c$), and waning of vaccine-induced immunity ($\gamma_z$). The left-most antigenic coordinate of the susceptible and vaccinated compartments represents individuals who have no natural infection memory.

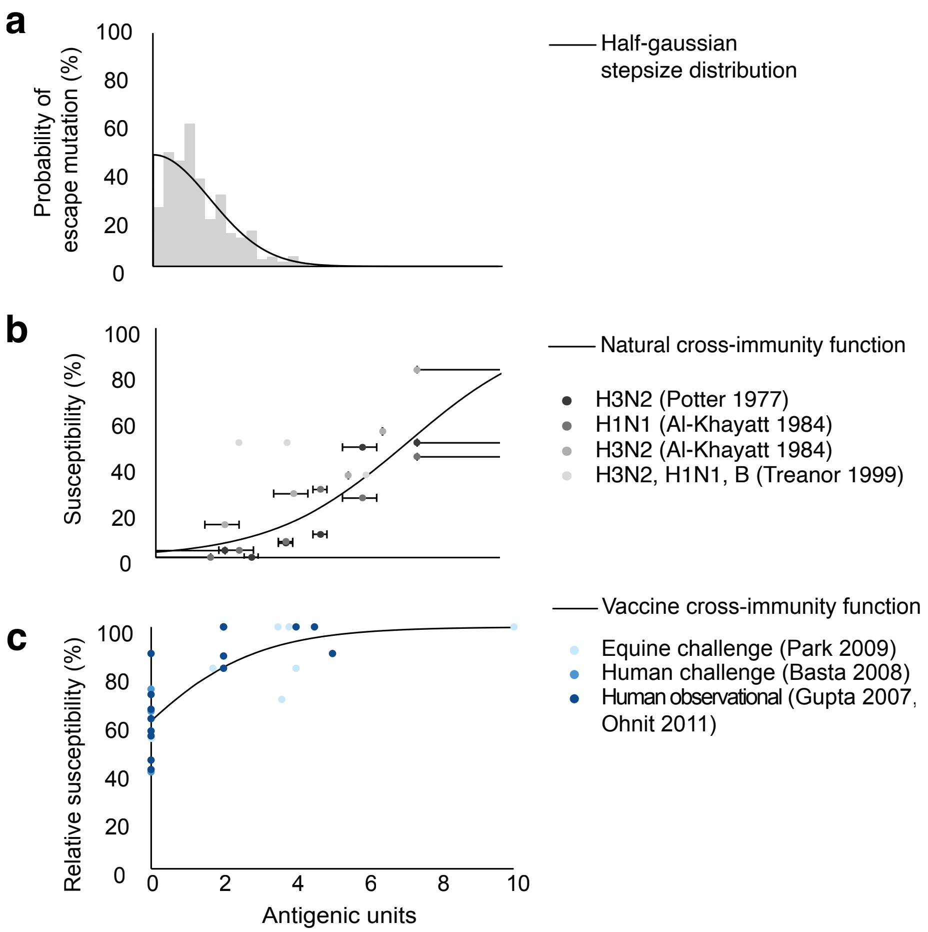


Figure S2: **Antigenic scales of mutations and cross-immunity for influenza A/H3N2. a** Stepsize distribution. The distribution is obtained from the average differences in logarithmically scaled HI titre values ($\log_2$HI) for 10,960 pairs of strains differing by a single amino acid substitution. The x-axis represents the antigenic distance in $\log_2$HI between the parent and the mutant strain. A half-gaussian probability distribution was fitted with a mean of 1.3. **b** Natural cross-immunity function. On the x-axis, data points and error bars show the mid-points and intervals of the antigenic distance between the memory and the infecting strain, calculated from $\log_2$HI titers. On the y-axis, data points represent the susceptibility, based on the percentage of individuals in each group who became infected. The solid line shows the logistic function, fitted to aggregated binary infection outcomes from the shown studies. **c** Vaccine cross-immunity function. Susceptibility is reduced by vaccination. Data points represent relative risks of infection or vaccine efficacies (y-axis) for varying distances between the vaccine and the infecting strain (x-axis). The solid line shows the logistic function, fitted to the aggregated data of the shown studies.

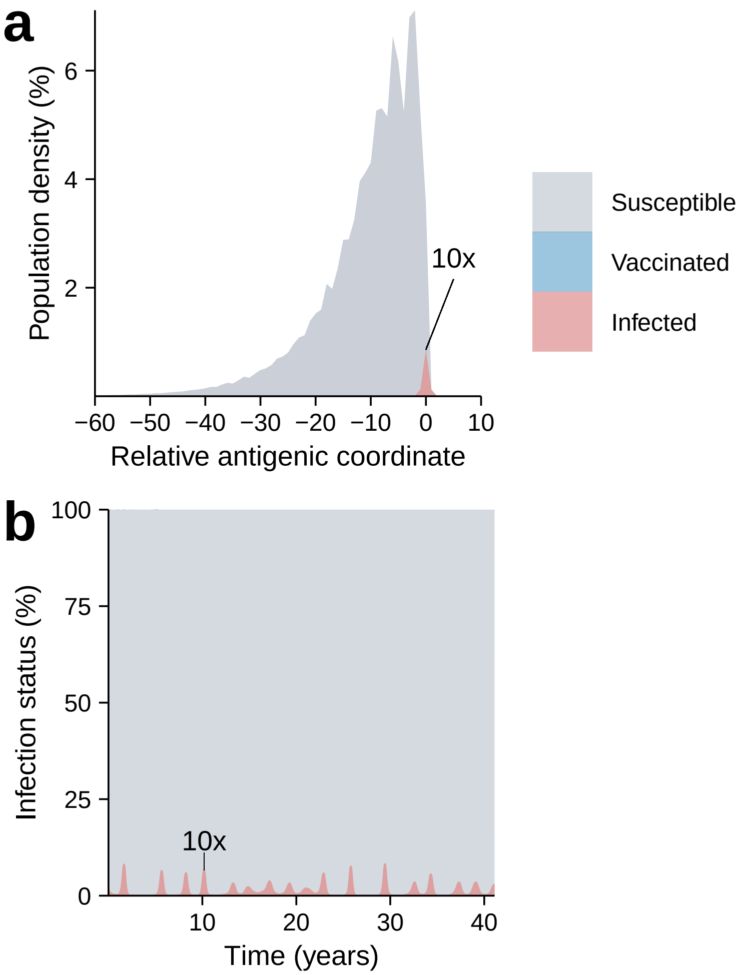


Figure S3: **Simulation outcomes from a single stochastic simulation run** for the main model without vaccination (Table 1, main text, for general parameters). Panel (**a**) shows the distributions of infected (red, 10× enlarged for clarity) and susceptible (grey) individuals according to the antigenic coordinates of the infecting strain or infection memory, centered at the mean of the infected wave at each time point and averaged over the simulation period. Panel (**b**) displays the infection prevalence over time.

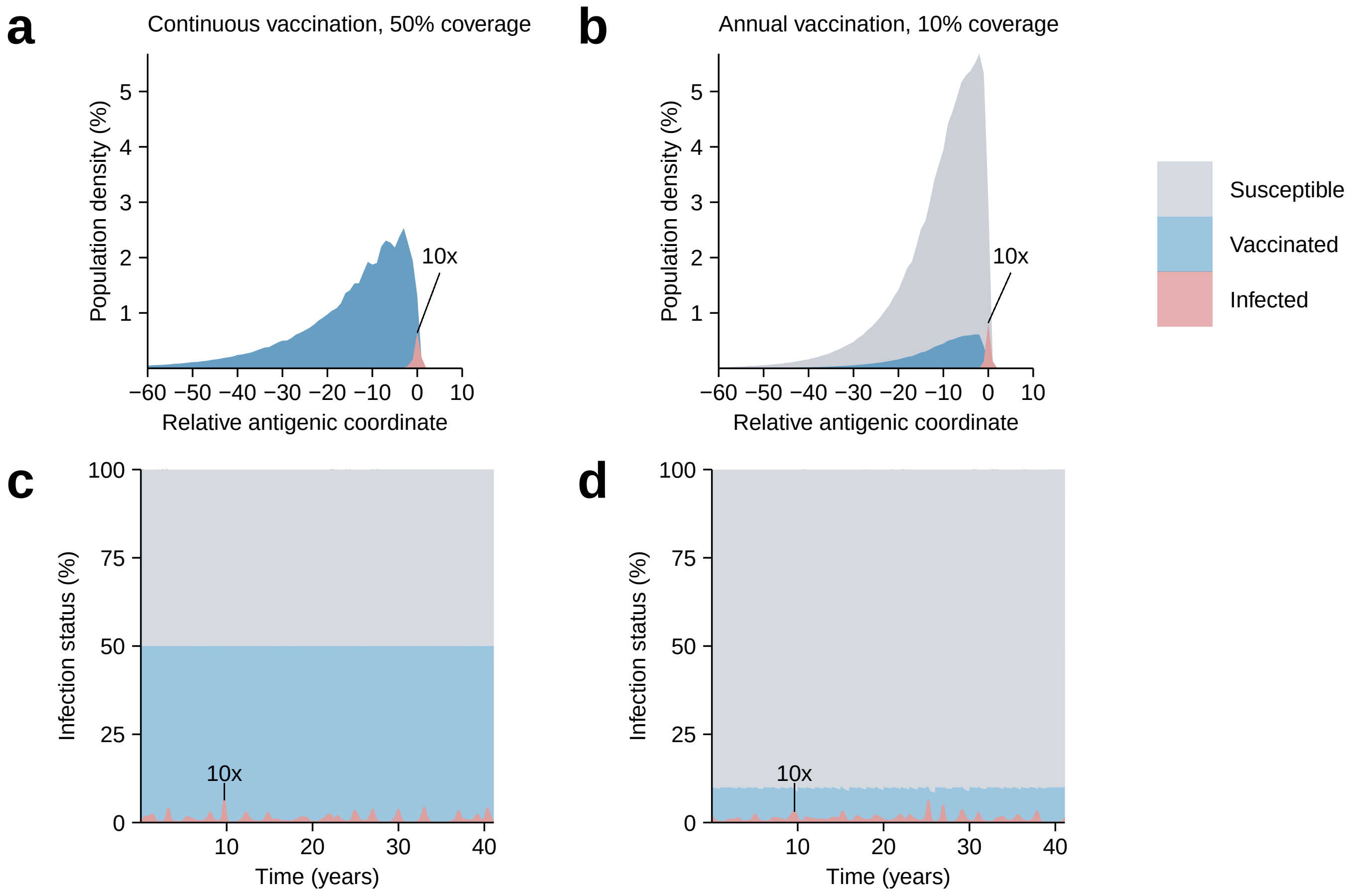


Figure S4: **Simulation outcomes from representative single stochastic runs** for: (**a,c**) the main model with continuous vaccination (Table 1, main text, for parameters), and (**b,d**) annual vaccination. Panels (**a,b**) show the distributions of infected (red, 10× enlarged for clarity), vaccinated (blue), and susceptible (grey) individuals across antigenic coordinates, centered at the mean of the infected wave at each time point and averaged over the simulation period, illustrating the movement and shape of the antigenic wave. Panels (**c,d**) display the percentages in each of the three compartments (infected, vaccinated, susceptible) over time.

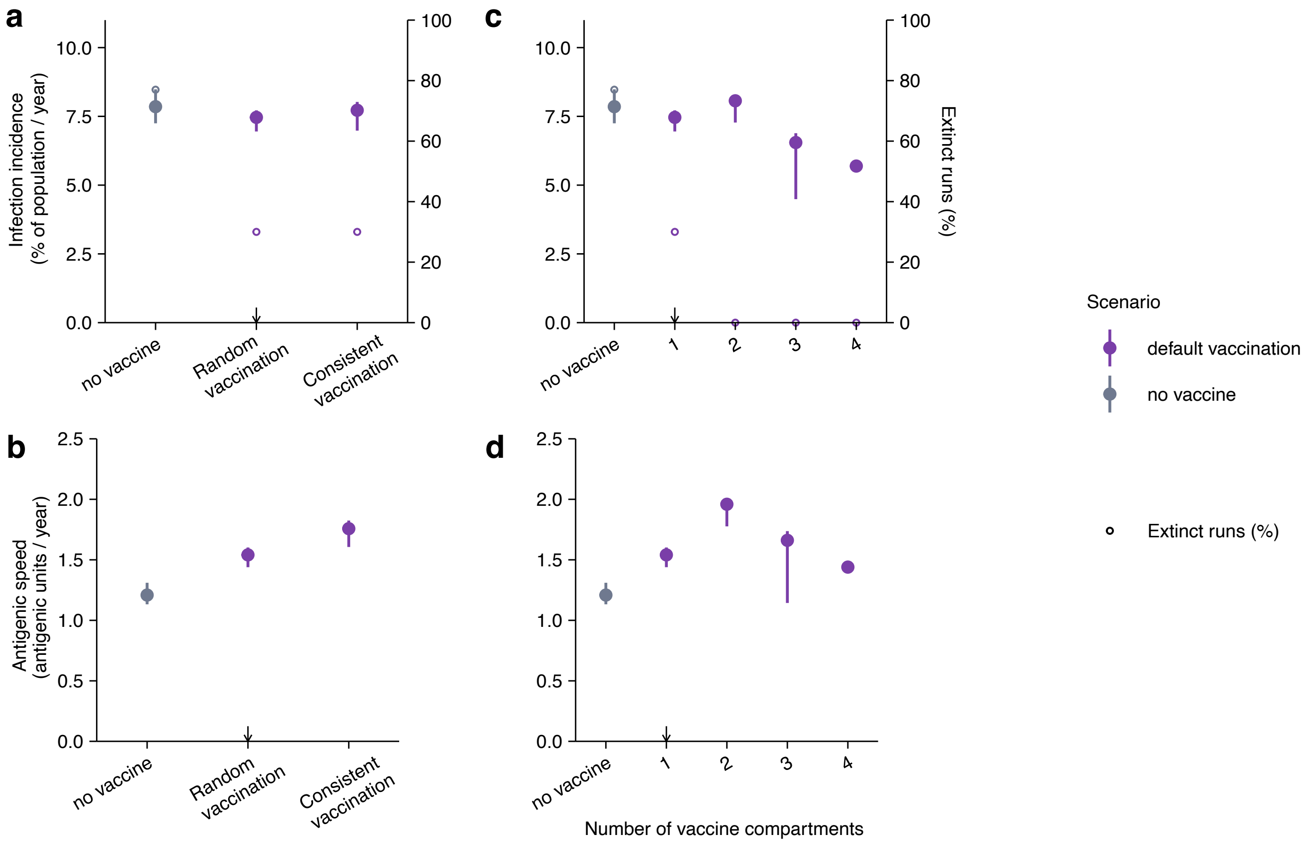


Figure S5: **Alternative models of vaccination.** Consistent vaccination (fixed subset vaccinated each round), multiple vaccination compartments (tracking recent vaccinations). The default vaccine scenario appears in purple, and the scenario without vaccination in grey. Panels (**a,c**) show annual infection incidence as a percentage of the population and panels (left y-axis) and the percentage of simulation runs resulting in pathogen extinction (open circles, right y-axis). Panels (**b,d**) show the speed of evolution in antigenic units per year. Outcomes were averaged over a 40-year simulation period. Small arrows on the x-axis indicate default scenarios.

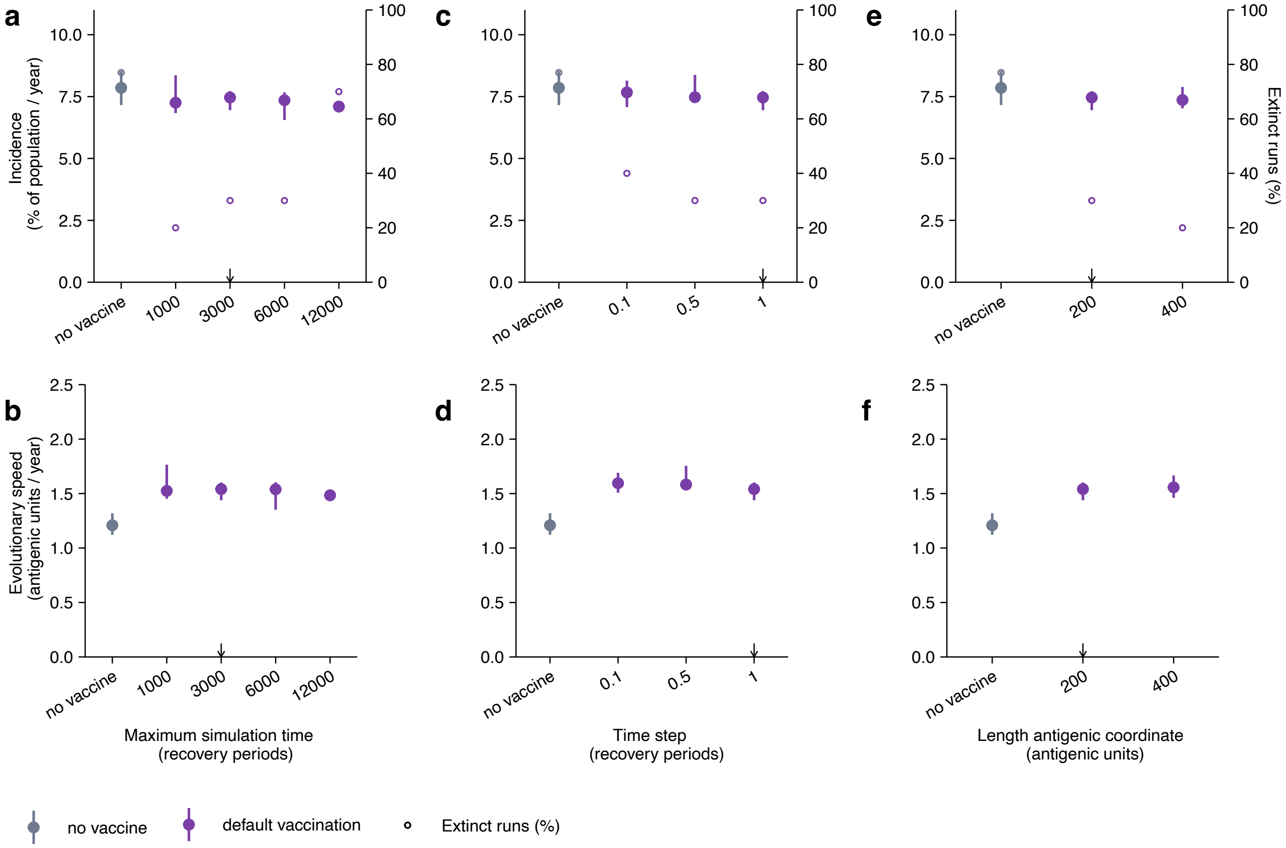


Figure S6: **The effect of simulation parameters**, duration of the simulation, the time step, and the length of the antigenic scale, on the outcomes. The default vaccine scenario appears in purple, and the scenario without vaccination in grey. Panels (**a,c,e**) show annual infection incidence as a percentage of the population (left y-axis) and the percentage of simulation runs resulting in pathogen extinction (open circles, right y-axis). Panels (**b,d,f**) show the speed of evolution in antigenic units per year, both averaged over a 40-year simulation period. Small arrows indicate default vaccination values, as specified in Table 1 in the main text. Outcomes were averaged over a 40-year simulation period. Small arrows indicate default values.

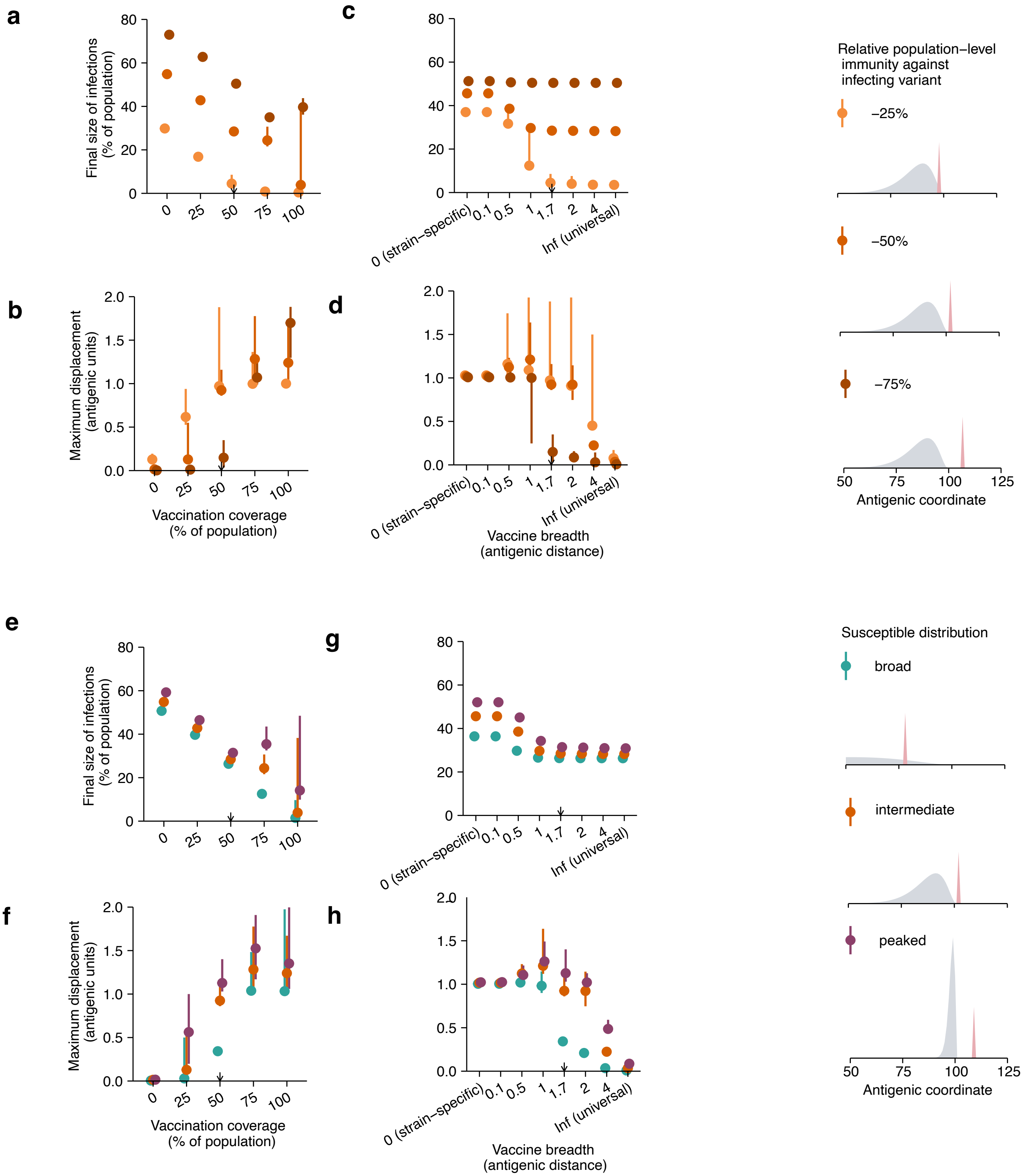


Figure S7: **Effect of vaccination on short-term evolution in a single outbreak.** Panels (**a,c,e,g**) show the final percentage of individuals that got infected during the outbreak, and panels (**b,d,f,h**) the maximum displacement of the mean of the infecting wave during the simulation period of 1 year. Small arrows indicate default vaccination values as specified in Table 1 in the main text. Outcomes are averaged over 40 years. Points represent the median and error bars the interquartile range across 10 stochastic simulation runs, regardless of whether they went extinct. Small arrows on the x-axis indicate default vaccination values (Table 1, main text). Here, vaccination was introduced only at the start of the simulations. The figures in the legend on the right show the initial distributions of the susceptible (grey) and infected (red, 100× enlarged for clarity) individuals.

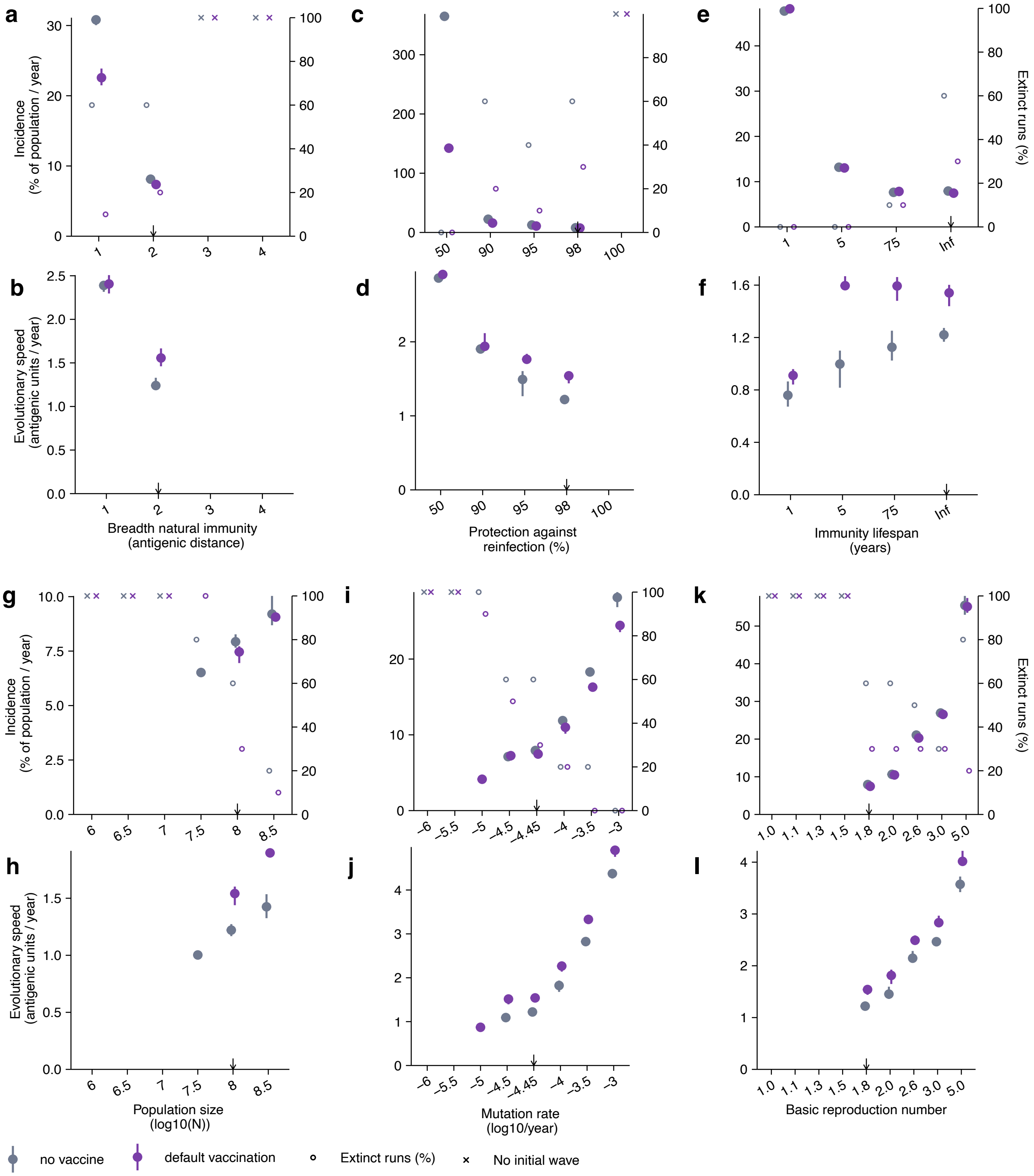


Figure S8: **Sensitivity to general parameters.** The effect of the reproduction number, population size, mutation rate, susceptibility to reinfection, and breadth of natural immunity on the epidemiological and evolutionary dynamics. The default vaccine scenario appears in purple, and the scenario without vaccination in grey, and a more extreme scenario of 100% coverage in red. Panels (**a,c,e,g,i,k**, left y-axis) show annual infection incidence as a percentage of the population and (right y-axis, open circles) the percentage of simulation runs resulting in pathogen extinction; crosses indicate cases where no stable traveling wave could be established. Panels (**b,d,f,h,j,l**) show the speed of antigenic evolution. Outcomes were averaged over a 40-year simulation period and compared to the no-vaccination scenario (grey). Points represent the median and error bars the interdecile range across 10 stochastic simulation runs that did not go extinct. Small arrows indicate default vaccination values as specified in Table 1 in the main text.

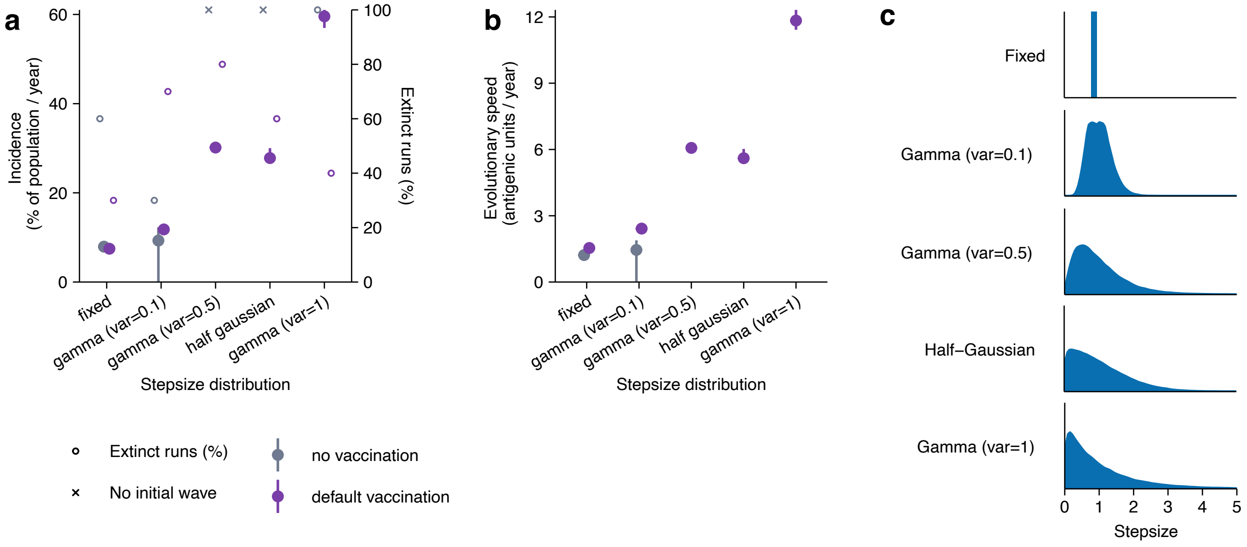


Figure S9: **Antigenic effect of mutations.** Simulation outcomes are shown for different distributions of antigenic mutation step sizes: fixed step size (default), gamma distributions with variances 0.1, 0.5 and 1 (the latter corresponding to an exponential distribution), and a half-Gaussian distribution. All distributions have a mean step size of 1 antigenic unit.
